# High-Density Surface Electromyography Reveals Shared Baseline Spatial Organization and Heterogeneous Fatigue Responses in Amyotrophic Lateral Sclerosis

**DOI:** 10.64898/2026.06.18.26355984

**Authors:** Ernesto H. Bedoy, Maia Brown, Michael A. Christofidis, Breanna Sullivan, Tawfiq Al-Lahham, Douglas Weber, Christi L. Kolarcik

**Affiliations:** Department of Mechanical Engineering Carnegie Mellon University, Pittsburgh, Pennsylvania, USA; University of Pittsburgh School of Medicine Pittsburgh, Pennsylvania, USA; Swanson School of Engineering University of Pittsburgh Pittsburgh, Pennsylvania, USA; Department of Neurology, University of Pittsburgh School of Medicine Pittsburgh, Pennsylvania, USA; Department of Pathology, University of Pittsburgh School of Medicine Pittsburgh, Pennsylvania, USA

**Author notes:** Corresponding Author: Christi L. Kolarcik, PhD, Department of Pathology, University of Pittsburgh School of Medicine, S701 Scaife Hall, 3550 Terrace Street, Pittsburgh, Pennsylvania 15213, USA.

**Keywords:** amyotrophic lateral sclerosis, high-density surface electromyography, spatial muscle activation, muscle fatigue, principal component analysis

## Abstract

**Introduction/Aims:** Amyotrophic lateral sclerosis (ALS) causes progressive motor neuron degeneration, denervation, collateral reinnervation, and altered motor unit organization. Clinical assessments track functional decline but provide limited information about the physiological remodeling that precedes or accompanies weakness. High-density surface electromyography (HD-sEMG) can noninvasively measure motor unit morphology, fatigue-related signal behavior, and spatial patterns of muscle activation.

**Methods:** We recorded HD-sEMG from the biceps brachii and tibialis anterior in participants with ALS and healthy controls during sustained isometric contractions at 30% and 50% maximum voluntary contraction. Features were extracted from four domains: fatigue dynamics, motor unit morphology, propagation, and spatial organization. Principal component analysis (PCA) was used to test whether the dominant HD-sEMG feature structure was shared or reorganized differently between groups at baseline and during fatigue.

**Results:** Baseline PCA showed highly similar HD-sEMG structure in healthy and ALS muscles. Baseline loading profiles were strongly spatial in both groups, with spatial features contributing 91.1% of loading weight in healthy observations and 89.9% in ALS observations. During fatigue, the composite did not significantly separate groups, but ALS showed a larger shift in loading structure and greater score variability than controls. The fatigue-change composite did not scale linearly with limb function. Exploratory binned analysis showed the greatest variability in the moderate impairment group.

**Discussion:** HD-sEMG captured strong spatial organization in both groups during baseline contraction. Sustained contraction exposed more variable ALS responses involving amplitude and spectral dynamics, rather than a single uniform fatigue pattern.

## INTRODUCTION

Amyotrophic lateral sclerosis (ALS) is a progressive neurodegenerative disease marked by muscle weakness, paralysis, and typical survival of about two to five years from diagnosis ^1^. Early in the disease course, motor neurons lose connections with muscle fibers at the neuromuscular junction, causing denervation ^2^. Surviving motor neurons partially compensate through collateral reinnervation, which can preserve strength temporarily and delay clinically obvious weakness ^3,4^.

Clinical measures such as the ALS Functional Rating Scale–Revised (ALSFRS-R) are essential for tracking function, but they do not directly measure the muscle-level remodeling that precedes or accompanies weakness ^5,6^. Needle electromyography (EMG) remains central to ALS diagnosis because it detects active and chronic denervation across body regions ^7^. However, it samples restricted muscle regions, depends on operator interpretation, and cannot quantify large-scale spatial recruitment patterns ^8,9^. These limitations create a need for noninvasive physiological measures that can sample broader muscle regions and track muscle-level changes in motor unit organization during disease monitoring and therapeutic studies ^10^.

High-density surface electromyography (HD-sEMG) addresses part of that gap by recording muscle activity across closely spaced electrode arrays ^11^. Unlike conventional single-channel recordings, HD-sEMG captures how activation is distributed across the muscle surface. It can quantify spatial activation patterns, estimate motor unit morphology through decomposition, and measure fatigue-related changes during sustained contraction ^12–15^. In ALS, previous work has examined motor unit firing behavior, motor unit action potential (MUAP) morphology, spectral fatigue measures, fasciculations, and amplitude- or spatial-based features ^16–19^. These studies support HD-sEMG as a promising physiological tool, but most analyses still treat feature domains separately. What remains unclear is which domains dominate when spatial, morphological, fatigue-related, and propagation features are analyzed together, and whether ALS changes that organization.

The present study used HD-sEMG recordings from the biceps brachii (BB) and tibialis anterior (TA) during sustained isometric contractions in participants with ALS and healthy controls. We extracted features reflecting spatial organization, motor unit morphology, fatigue dynamics, and propagation, then used principal component analysis (PCA) to identify dominant patterns of variation. We analyzed features from the initial steady portion of contraction separately from the fatigue-related changes across the contraction to determine whether dominant HD-sEMG structure was present at baseline, changed during sustained effort, or differed between participants with ALS and healthy controls. We hypothesized that ALS would alter the multifeature organization of HD-sEMG signals, and that baseline and fatigue-change analyses could reveal different aspects of neuromuscular remodeling.

## METHODS

### A. Participants

We recruited 20 participants: 8 with ALS and 12 healthy controls. Participants with ALS met revised El Escorial diagnostic criteria. Participants were excluded for severe cognitive or physical impairment, recent neurological or musculoskeletal disorders unrelated to ALS, or conditions that could interfere with EMG recordings. The initial visit was used for cross-sectional analyses. Follow-up visits and exclusions are described in the Supplementary Results. The study was approved by the University of Pittsburgh Institutional Review Board (Protocol #STUDY22120029), and written informed consent was obtained before each visit.

### B. Experimental Protocol

HD-sEMG was recorded from the TA and BB to sample lower- and upper-limb muscles and account for regional heterogeneity in ALS (**Figure 1**). Participants performed isometric biceps flexion and ankle dorsiflexion with their dominant limb, except H06 and H07, who used the left limb, against a handheld dynamometer (Hoggan MicroFET2, Salt Lake City, UT). Participants first generated maximum voluntary contractions (MVCs), which were used to normalize target force. They then performed sustained contractions at 30% and 50% MVC for up to five minutes. Two force levels were selected to preferentially engage different motor unit populations across contraction intensities. We implemented a one-minute rest period between contractions for recovery. Because participants self-reported fatigue one minute before stopping, contraction duration was treated as an indirect fatigue measure. For feature analysis, contractions were divided into an Early window, defined as the first 10 s of steady contraction, and a Late window, defined as the final 10 s.

**Figure 1.**
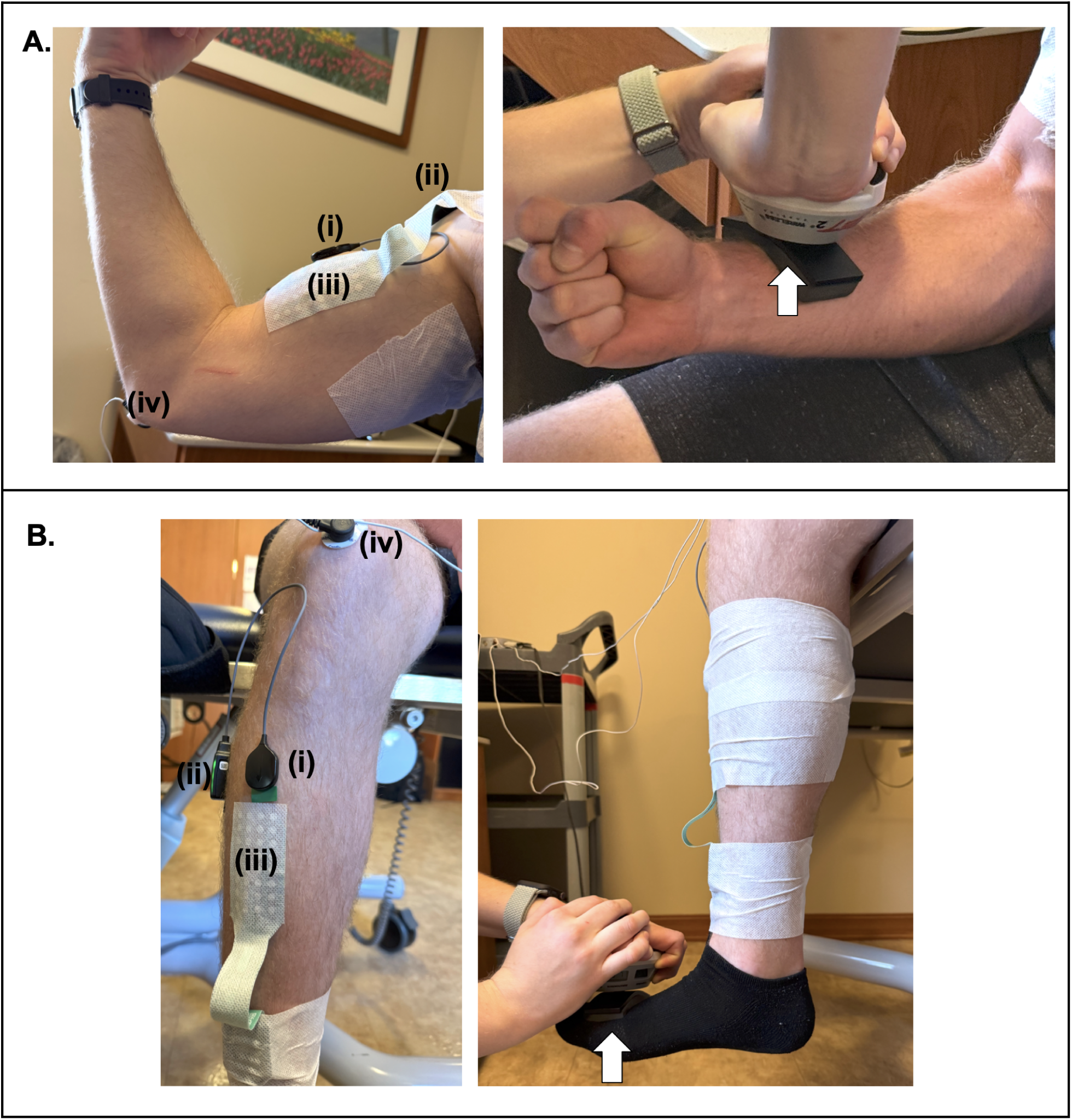
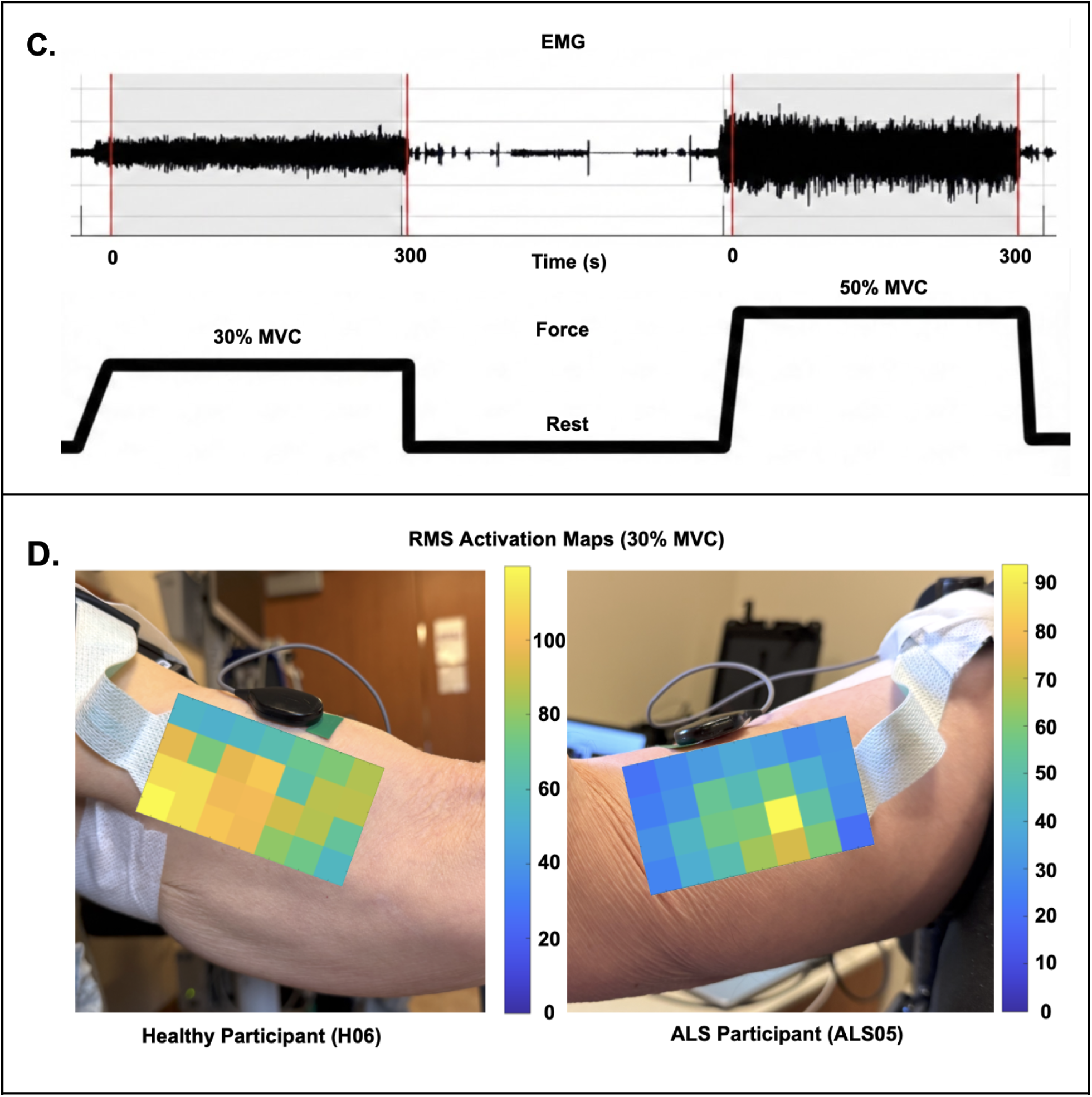
Experimental setup, contraction protocol, and representative spatial activation maps. **A**, HD-sEMG instrumentation and task configuration for the biceps brachii (BB). **B,** HD-sEMG instrumentation and task configuration for the tibialis anterior (TA). Electrode placements are shown for both the decomposition and spatial HD-sEMG systems, including the decomposition HD-sEMG sensor (i) and reference (ii), spatial HD-sEMG grid (iii), and spatial reference (iv). Participants performed isometric biceps flexion and ankle dorsiflexion against a handheld dynamometer, with arrows indicating the direction of applied force. **C,** Representative single-channel differentiated HD-sEMG and force traces during sustained contractions at 30% and 50% maximum voluntary contraction (MVC), separated by a rest period. Red lines on the EMG trace mark the start and end of the contraction interval shown; contraction duration varied across participants. **D,** Representative RMS activation maps from the biceps brachii during a 30% MVC contraction in a healthy participant (H06; left) and a participant with ALS (ALS05; right). The healthy example shows more distributed activation across the grid, whereas the ALS example shows a more focal region of higher activation intensity. These maps illustrate the spatial information captured by HD-sEMG and quantified in the spatial feature analyses.

### C. HD-sEMG Acquisition

Skin was prepared with abrasive gel (Lemon Prep, Mavidon, Flat Rock, NC) to reduce impedance. HD-sEMG was recorded using two complementary systems: a wireless decomposition sensor for motor unit decomposition (Trigno Galileo, Delsys Inc., Natick, MA, USA) and a 32-channel spatial grid for distributed muscle activation mapping (SAGA 32+, TMSi, Oldenzaal, Netherlands). Decomposition recordings were used to estimate motor unit firing behavior and MUAP morphology. Spatial grid recordings were used to quantify root-mean-square (RMS) amplitude, spectral features, and spatial activation patterns. Detailed acquisition parameters, preprocessing steps, and quality-control criteria are provided in the Supplementary Methods.

### D. Feature Extraction

Features were extracted across four physiological domains: fatigue dynamics, motor unit morphology, propagation, and spatial organization. Fatigue-dynamics features captured amplitude, spectral power, and motor unit firing behavior. Morphology features captured MUAP amplitude, duration, and waveform complexity. Propagation features captured conduction-related timing and waveform similarity. Spatial features captured how RMS activation was distributed across the electrode grid. These domains were selected because ALS can affect the number, size, firing behavior, and spatial territory of surviving motor units. Detailed feature definitions are provided in the Supplementary Methods.

### E. Limb Function Assessment

Limb function was treated as a continuous variable using a self-reported 10-point scale, where 1 indicated complete paralysis and 10 indicated normal strength. Only ALS observations were included in analyses relating electrophysiological features to limb function.

### F. Data Processing and PCA

Features were aggregated at the observation level, defined as Subject × Visit × Muscle × Target Force Level. Two matrices were generated. The Early-window matrix used feature values from the first 10 s of steady contraction. The fatigue-change matrix expressed each feature as the percent change from Early to Late. Features were standardized before PCA so that differences in feature scale did not dominate the components.

Two PCA approaches were used. Group-specific PCAs were run separately for Healthy Early, ALS Early, Healthy fatigue-change, and ALS fatigue-change observations. These analyses identified which feature domains dominated the internal structure of each group and condition. Common-axis PCAs were then run across the full cohort to project Healthy and ALS observations onto the same component and compare PC1 scores between groups. Additional details on normalization, PCA implementation, domain-restricted analyses, and propagation-feature handling are provided in the Supplementary Methods.

### G. Statistical Analysis

Linear mixed-effects models tested common-axis group differences in PC1 score, with group as the primary fixed effect and subject identity as a random intercept. Contraction force and muscle were included as fixed effects when the model structure allowed. Relationships between the fatigue-change composite and limb function were tested in ALS observations using mixed-effects models. Severity-bin and longitudinal analyses were exploratory and are described in the Supplementary Methods. Statistical significance was defined as p < 0.05, and analyses were performed in MATLAB (MathWorks, Natick, MA, USA).

## RESULTS

### A. Cohort Characteristics

The final dataset included baseline recordings from 17 participants: 7 with ALS and 10 healthy controls. Age (ALS: 63.1 ± 5.1 years; Healthy: 63.0 ± 15.8 years; Welch’s t-test, p = 0.979) and sex distribution (57% male in ALS; 60% male in Healthy) were similar between groups. Disease duration since diagnosis ranged from 4 to more than 180 months, and self-reported clinical scores ranged from 1.5 to 10 across affected limbs (**Table 1**). The PCA dataset included 96 collapsed observations: 52 Healthy and 44 ALS observations. Common-axis group comparisons included 52 Healthy and 41 ALS observations after feature-level filtering and PCA row inclusion. Data exclusions, MVC strength, contraction duration, decomposition yield, and longitudinal stability are reported in the Supplementary Results.

**Table 1.** Disease duration and self-reported clinical muscle function. Approximate disease duration since diagnosis, reported in months, and self-rated clinical scores at each visit for participants with ALS. Functional ratings ranged from 1 (complete paralysis) to 10 (normal strength). Commas separate values across multiple visits. BB = biceps brachii; TA = tibialis anterior.

| Subject ID | Disease Duration<br>(months) | Clinical Score<br>(BB) | Clinical Score (TA) |
| --- | --- | --- | --- |
| ALS01 | > 180, + 7, + 4 | 9, 9, 9 | 1.5, 1.5, 1.5 |
| ALS02 | ~19, + 3, + 2 | 8, 8, 7 | 10, 10, 10 |
| ALS03 | ~49 | 4 | 5 |
| ALS04 | ~4 | 9 | 9 |
| ALS05 | ~12.5 | 7 | 7 |
| ALS06 | 9 | N/A | N/A |
| ALS07 | 43 | 6 | 9 |

### B. Baseline HD-sEMG Loading Structure is Shared Between Groups and Dominated by Spatial Features

Early-window PCA showed that spatial organization was the dominant source of HD-sEMG variation in both groups (**Figure 2; Table S1**). In Healthy observations, PC1 explained 45.20% of variance and was driven primarily by cluster index, heterogeneity, peak-to-median ratio, active area, spatial entropy, RMS variance, and dispersion. Spatial features accounted for 91.1% of PC1 loading weight. In ALS observations, PC1 explained 38.97% of variance and was also spatially dominated, with spatial features accounting for 89.9% of loading weight. The Early-window loading profiles were highly similar between groups (cosine similarity = 0.949; Spearman rho = 0.933; **Table S2**). Baseline HD-sEMG therefore had a strongly spatial structure in both Healthy and ALS muscle.

**Figure 2.**
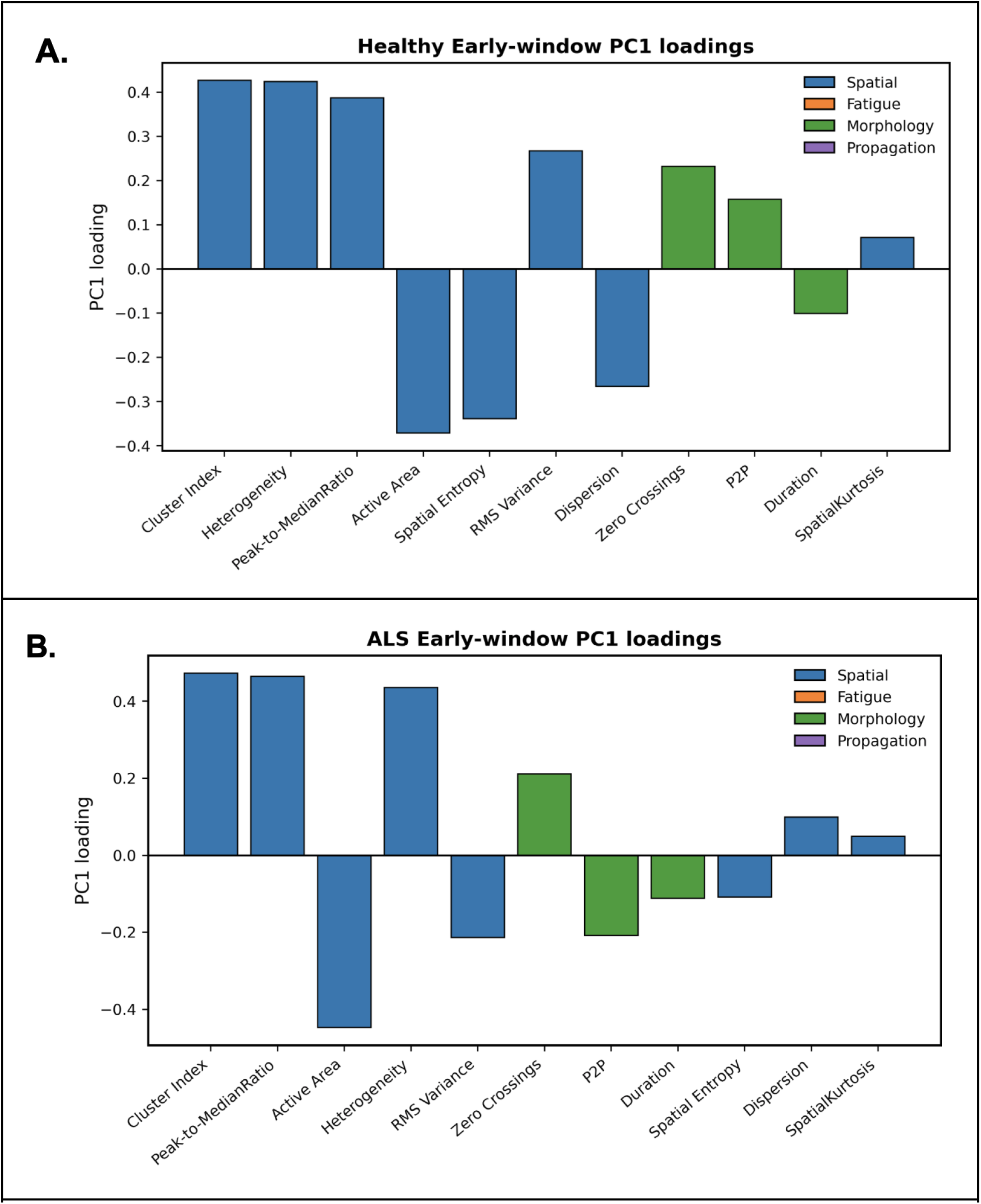
Early-window PC1 loading structure in healthy and ALS muscle. **A**, Group-specific Early-window PCA loadings for Healthy observations. **B,** Group-specific Early-window PCA loadings for ALS observations. In both groups, PC1 was dominated by spatial features, including cluster index, heterogeneity, peak-to-median ratio, active area, spatial entropy, RMS variance, and dispersion. Spatial features accounted for 91.1% of PC1 loading weight in Healthy observations and 89.9% in ALS observations, indicating that baseline HD-sEMG structure was strongly spatial in both groups. P2P = peak-to-peak amplitude.

Because spatial features dominated baseline loadings, we tested whether they also separated ALS from controls. They did not. The spatial-only Early-window common-axis PCA showed no significant group effect (ALS minus Healthy: β = 0.341, p = 0.458; **Figure 3A; Table S3**), and no individual spatial feature reached significance (**Figure 3B; Table S4**). Dispersion showed the strongest trend toward lower values in ALS (β = -0.765 Healthy SD units, p = 0.057). These results indicate that spatial organization was the dominant baseline HD-sEMG structure, but it was not a standalone group-separating signal in this cohort.

**Figure 3.**
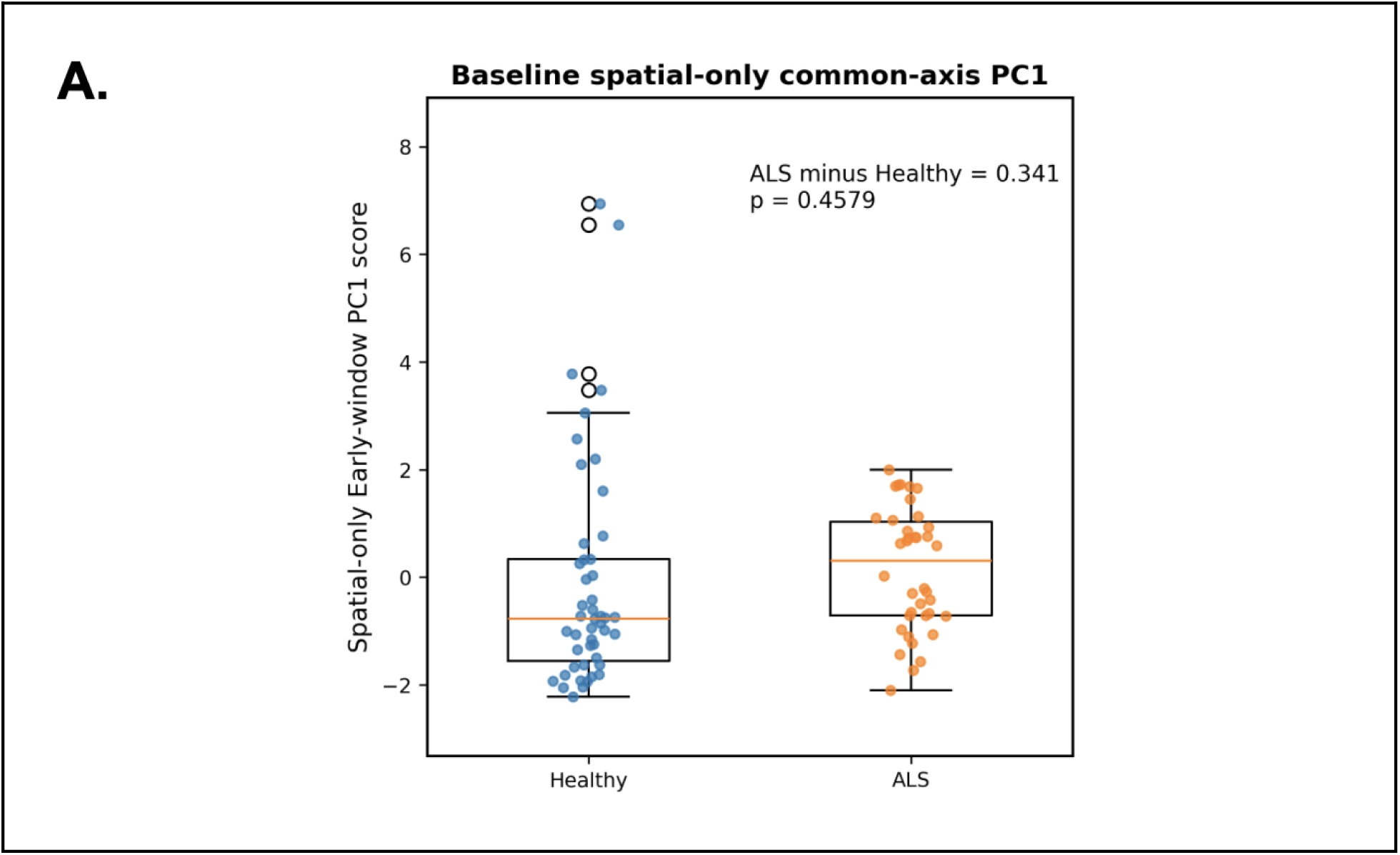
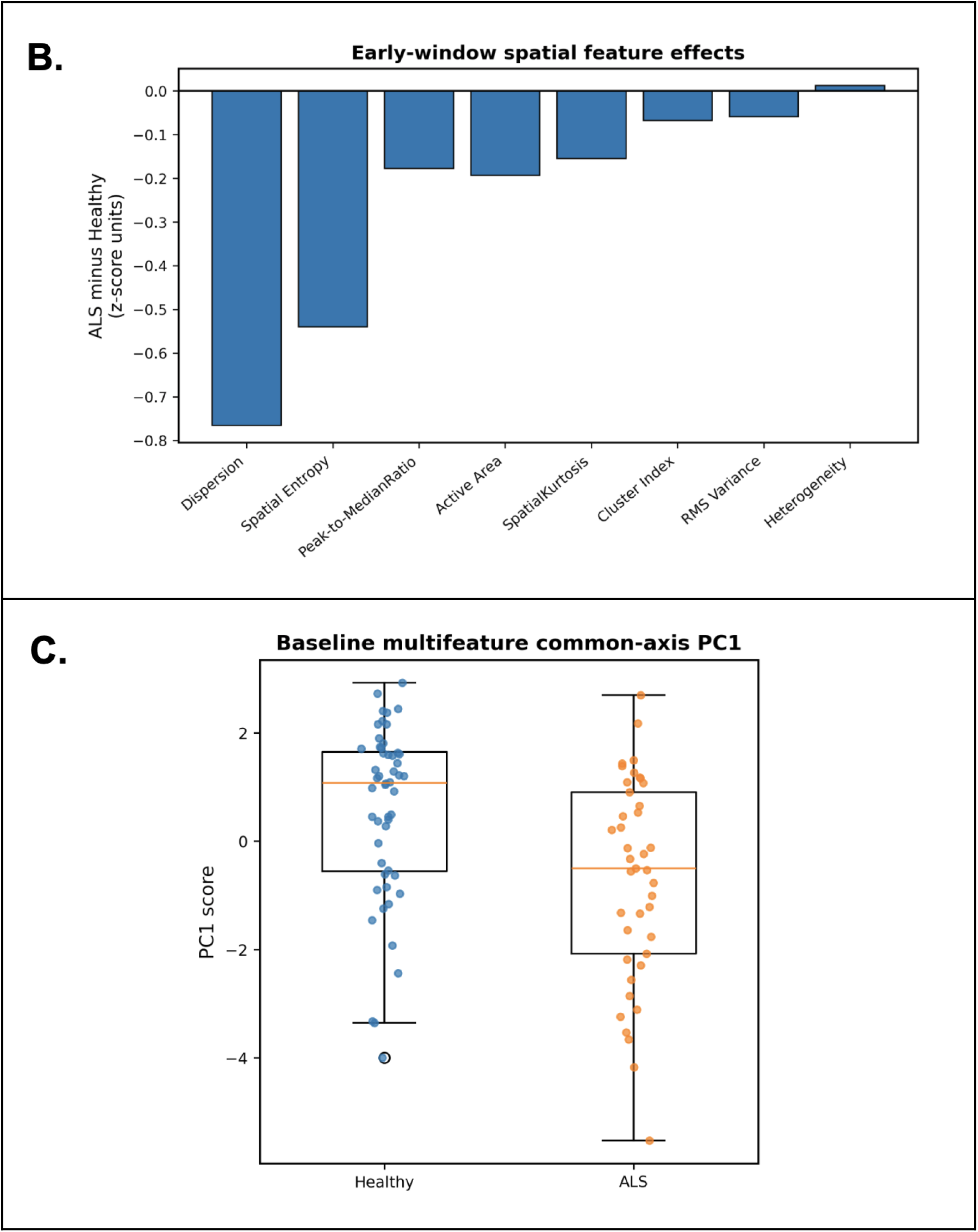
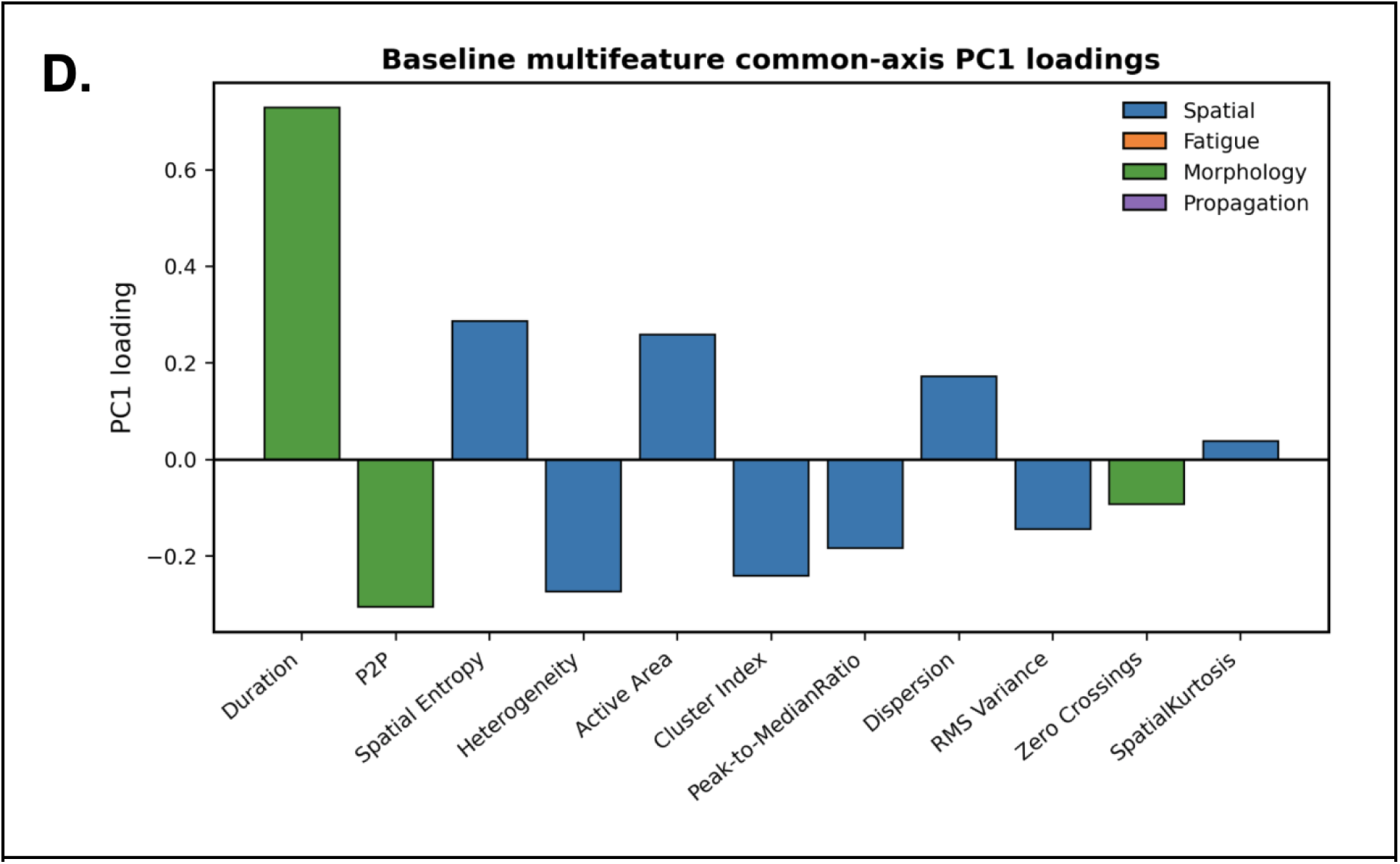
Baseline spatial-only and multifeature PCA comparisons. **A–B**, Early-window spatial-only common-axis PCA. **A,** Distribution of PC1 scores in Healthy and ALS observations. The spatial-only PC1 explained 43.60% of total variance and did not differ significantly between groups (ALS minus Healthy = 0.341, p = 0.4579). **B,** Individual Early-window spatial feature estimates, reported as ALS minus Healthy in Healthy-referenced z-score units. Negative values indicate lower values in ALS. No individual spatial feature reached statistical significance, although dispersion showed the strongest trend toward lower values in ALS. **C,** All-feature Early-window common-axis PCA. The multifeature PC1 explained 30.36% of total variance and separated ALS from Healthy observations, with mean scores of 0.553 ± 1.623 in Healthy and -0.702 ± 1.891 in ALS (ALS minus Healthy = -1.372, p = 0.0016). **D,** PC1 loadings for the all-feature Early-window common-axis PCA color-coded by feature domain. The baseline multifeature axis was defined by both morphology and spatial features, with MUAP duration contributing the largest loading. P2P = peak-to-peak amplitude.

### C. Baseline HD-sEMG Composite Separates ALS from Healthy Controls

The all-feature Early-window common-axis PCA separated ALS from Healthy controls (**Figure 3C; Table S3**). PC1 explained 30.36% of variance. Mean PC1 score was 0.553 ± 1.623 in Healthy observations and -0.702 ± 1.891 in ALS observations, with a significant group effect (ALS minus Healthy: β = -1.372, p = 0.0016). The loading structure showed why this result differed from the spatial-only analysis. MUAP duration was the largest contributor to the common-axis PC1 (loading = 0.729; squared loading = 0.531), followed by peak-to-peak amplitude and several spatial features, including entropy, heterogeneity, active area, cluster index, peak-to-median ratio, dispersion, and RMS variance (**Figure 3D**). Baseline group separation therefore required information from both morphology and spatial activation.

### D. Fatigue Reorganizes the ALS Feature Profile More than the Healthy Profile

Fatigue-change PCA produced a different pattern (**Figure 4; Table S1**). In Healthy observations, PC1 explained 29.82% of variance and remained mostly spatial. Spatial features accounted for 80.5% of loading weight and fatigue-dynamics features accounted for 16.5%. In ALS observations, PC1 explained 31.74% of variance, but the loading structure was more mixed. Spatial features accounted for 57.9% of loading weight, fatigue-dynamics features increased to 34.4%, and low-frequency power, RMS amplitude, and high-frequency power were among the strongest fatigue-change loadings.

**Figure 4.**
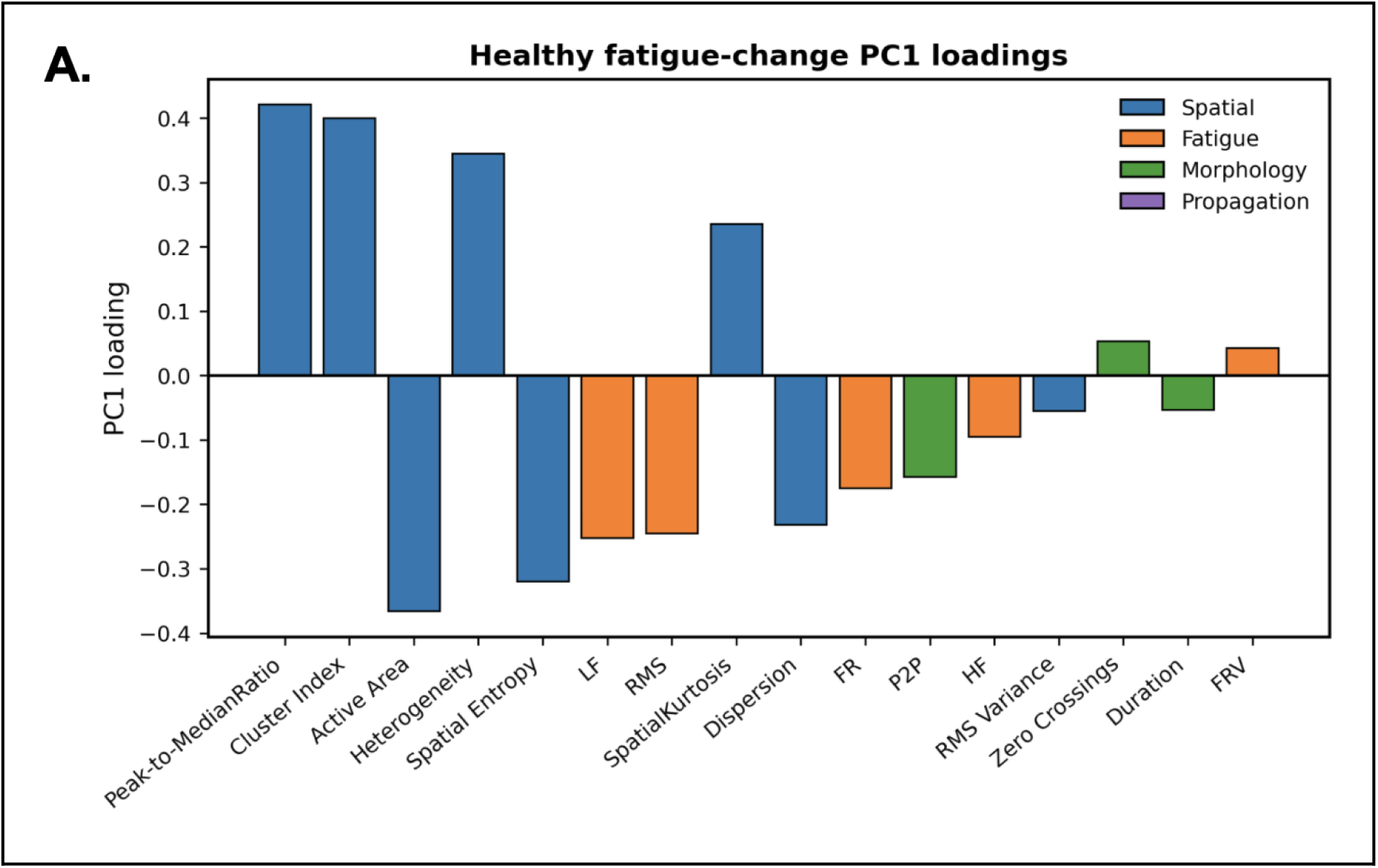
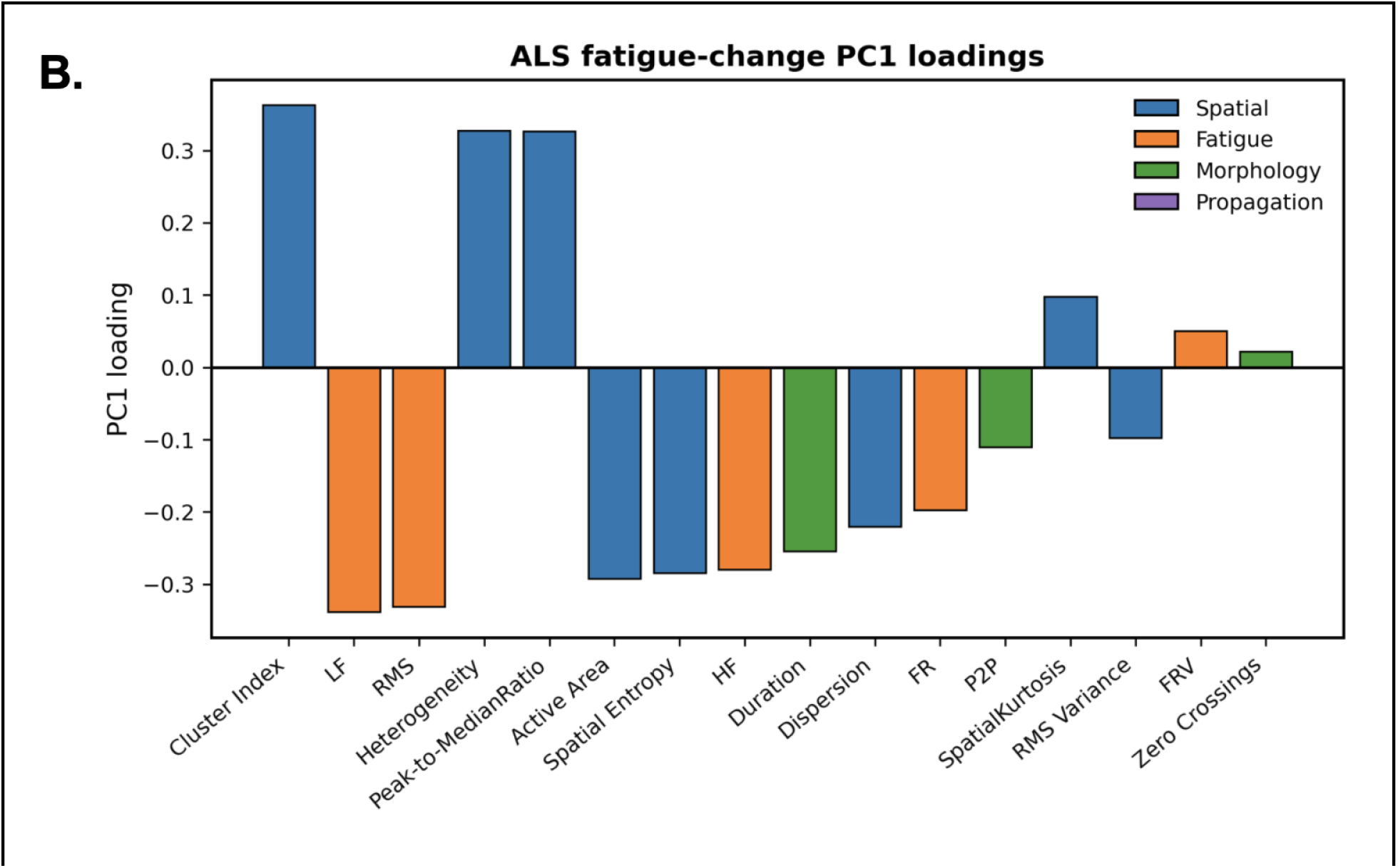
Fatigue-change PC1 loadings in healthy and ALS muscle. **A**, PC1 loadings for the group-specific Healthy fatigue-change PCA. PC1 explained 29.82% of total variance. Spatial features accounted for 80.5% of PC1 loading weight and fatigue features accounted for 16.5%, indicating that the healthy fatigue response remained mostly spatial. **B,** PC1 loadings for the group-specific ALS fatigue-change PCA. PC1 explained 31.74% of total variance. Spatial features accounted for 57.9% of PC1 loading weight, whereas fatigue features accounted for 34.4%, indicating a more mixed fatigue-related feature structure in ALS. Bars are color-coded by feature domain. LF = low-frequency power; HF = high-frequency power; FR = firing rate; FRV = firing rate variability; P2P = peak-to-peak amplitude

The Early-to-fatigue-change shift was larger in ALS than in controls (**Table S2**). Healthy Early and Healthy fatigue-change loading patterns showed moderate similarity (cosine similarity = 0.860; Spearman rho = 0.591). ALS Early and ALS fatigue-change patterns were less similar, especially by rank order (cosine similarity = 0.731; Spearman rho = 0.188). Sustained contraction preserved much of the Healthy loading structure but changed the ALS structure more substantially.

Despite this reorganization, the common-axis fatigue-change PCA did not significantly separate groups (**Figure 5; Table S3**). PC1 explained 33.04% of variance. Mean PC1 score was 0.284 ± 1.938 in Healthy observations and -0.361 ± 3.612 in ALS observations, with no significant group effect (ALS minus Healthy: β = -0.614, p = 0.2837). Although the fatigue-change composite did not significantly separate groups, the wider distribution of ALS scores indicates that sustained contraction produced a more variable physiological response in ALS muscles.

**Figure 5.**
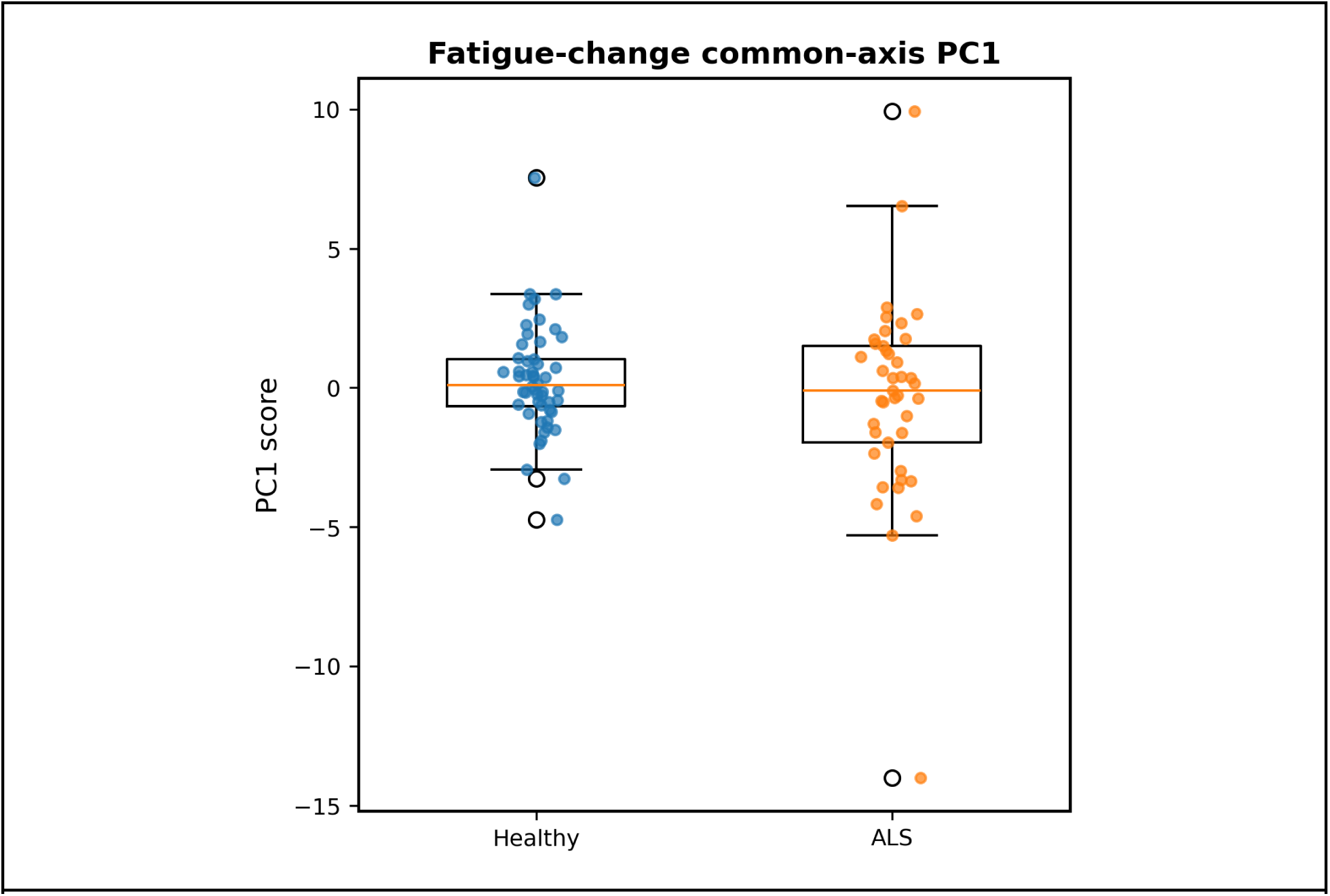
Fatigue-change common-axis group comparison. Distribution of PC1 scores from the all-feature fatigue-change common-axis PCA in Healthy and ALS observations. PC1 explained 33.04% of total variance. Healthy observations had a mean score of 0.284 ± 1.938, whereas ALS observations had a mean score of -0.361 ± 3.612. The mixed-effects model did not show a significant group effect (ALS minus Healthy = -0.614, p = 0.2837), but ALS observations showed greater spread.

### E. Clinical Coupling and Disease Severity

The all-feature fatigue-change common-axis PC1 score did not scale linearly with limb function in ALS (β = 0.021, p = 0.913; **Figure 6A**). Pearson and Spearman correlations were also negligible (r = 0.017, p = 0.915; rho = -0.071, p = 0.646). Severity-bin analysis showed broad overlap across groups, but the moderate impairment group had greater variability and a modest shift toward more negative composite values (**Figure 6B**). These results argue against a simple linear relationship between fatigue-related HD-sEMG change and clinical weakness. Exploratory longitudinal analyses are reported in the Supplementary Results.

**Figure 6.**
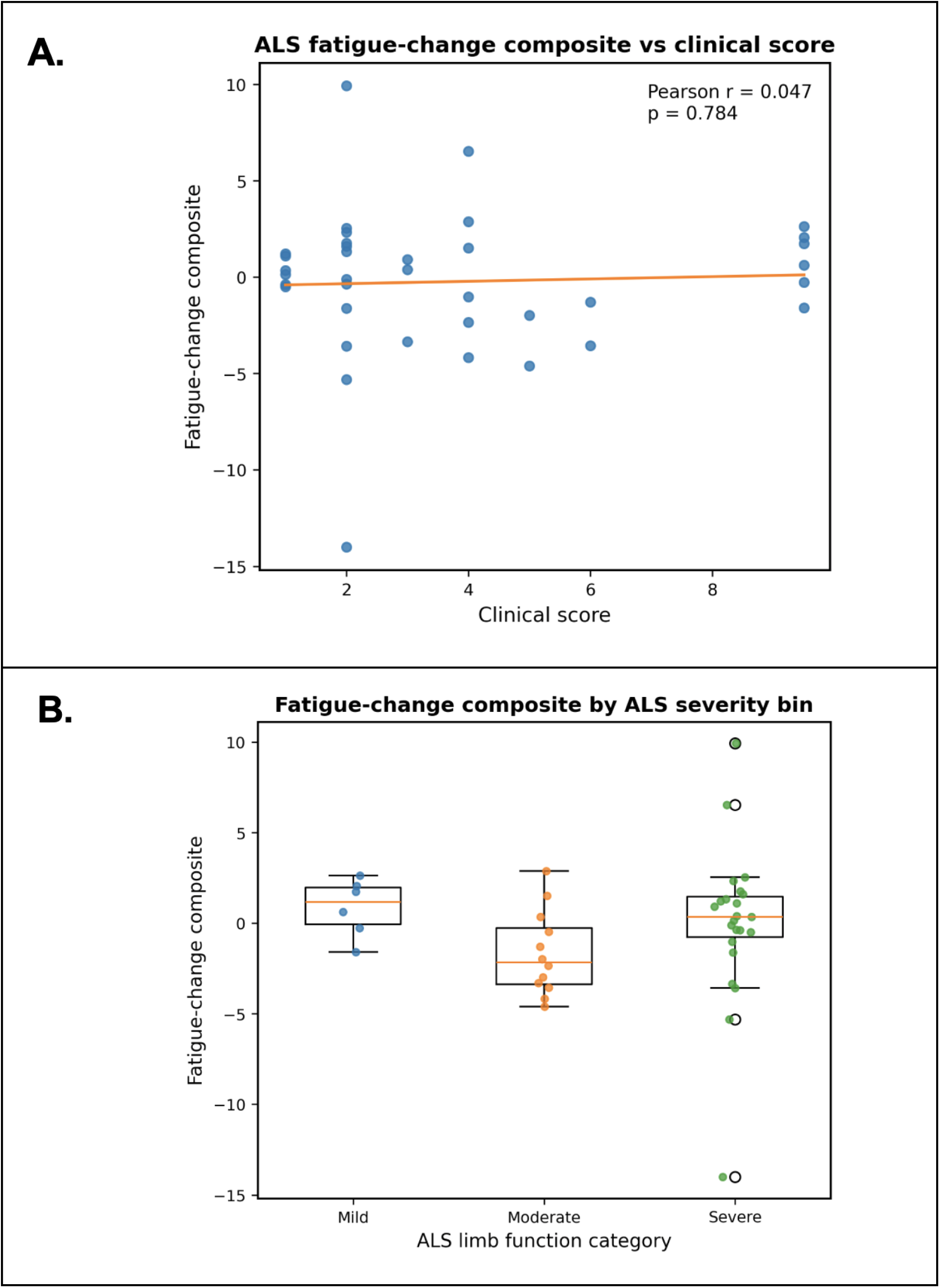
Fatigue-change composite in relation to clinical function and ALS severity. **A**, Relationship between the all-feature fatigue-change common-axis PC1 score and self-reported limb function score in ALS observations. The fitted line shows that the fatigue-change composite did not scale linearly with clinical function in this cohort. **B,** Distribution of the all-feature fatigue-change common-axis PC1 score across ALS severity bins defined from limb function scores. Composite values overlapped across severity groups, but the moderate impairment group showed greater variability and a modest shift toward more negative values. Together, these findings suggest that fatigue-related HD-sEMG responses vary across ALS observations rather than changing uniformly with clinical severity.

## Discussion

This study produced two main findings. First, baseline HD-sEMG structure was strongly spatial and highly similar in healthy and ALS muscles. Spatial features dominated Early-window PC1 in both groups, but spatial-only analyses did not separate participants with ALS from healthy controls. Second, sustained contraction changed the ALS feature profile more than the Healthy profile. Fatigue shifted ALS loadings toward a mixed spatial, RMS, and spectral structure and increased score variability. Thus, baseline HD-sEMG captured shared spatial organization, whereas fatigue exposed heterogeneous ALS responses under sustained demand.

### A. Shared Baseline Spatial Organization

The within-group PCAs showed that spatial features captured most baseline HD-sEMG structure in both groups. During the early steady contraction window, the largest source of variation was how activation was distributed across the muscle surface. Spatial entropy, dispersion, clustering, heterogeneity, active area, RMS variance, and peak-to-median ratio describe related but distinct features of this distribution, including spread, focality, and unevenness of activation ^20–24^. Their dominance means that early HD-sEMG recordings are especially sensitive to the organization of recruited motor unit territories and regional muscle activation.

The high similarity between Healthy and ALS Early-window loadings means that this dominant spatial structure was largely preserved in ALS. That should not be interpreted as normal ALS physiology. Rather, it suggests that during a brief steady contraction, ALS muscles could still organize activity along a spatial axis similar to controls. A likely explanation is compensation. Remaining motor units, including enlarged reinnervated units, may be sufficient to maintain the target force over a short window ^3,25,26^. In that setting, spatial HD-sEMG captures how activation is organized, but it may not distinguish whether the organization is produced by a normal motor unit pool or by a remodeled pool that can still meet the task demand.

### B. Baseline Multifeature Differences

The all-feature Early-window common-axis PCA did separate ALS from controls. This is an important distinction because it shows that the dominant structure and the group-separating structure were not identical. Spatial features dominated the baseline signal, but group separation required morphology as well. MUAP duration contributed most strongly, with additional contributions from peak-to-peak amplitude and spatial features. We interpret this baseline axis as a multifeature signature of chronic motor unit remodeling. Denervation and collateral reinnervation can alter MUAP morphology, while changes in motor unit territories and recruitment can shape the spatial activation map ^24,26,27^. The present data do not isolate one mechanism, but they show that baseline HD-sEMG carries ALS-related information when morphology and spatial organization are analyzed together.

### C. Fatigue Reveals Heterogeneous ALS Responses

Fatigue changed the ALS feature structure in a way that baseline analysis did not. In controls, fatigue-change PC1 remained mostly spatial and composite scores were relatively compact. In ALS, fatigue-change PC1 became more mixed, with larger contributions from RMS amplitude and spectral power, and the composite scores spread widely. Rather than producing a single shared ALS-control shift, sustained contraction revealed greater variability across ALS observations.

We interpret sustained contraction as a physiological stressor on a remodeled motor unit pool. Fatigue increases demand on motor unit reserve, firing stability, and muscle fiber conduction properties ^15,19,28,29^. In ALS, those demands are imposed on muscles with variable denervation, reinnervation, residual strength, and regional involvement. Some muscles may compensate by increasing amplitude or recruitment, whereas others may show spectral or spatial changes as available motor unit pools fatigue or fail. This may explain why amplitude and spectral features became more prominent in ALS without producing a uniform group-level shift ^3,15,19,25,26,29–31^.

This interpretation also helps explain why fatigue may still be useful, even though it did not more clearly distinguish participants with ALS from healthy controls in this cohort. A uniform diagnostic marker would shift ALS observations in the same direction. That is not what we saw. Instead, sustained contraction uncovered a less stable feature structure across ALS observations. For a heterogeneous disease such as ALS, that variability may be biologically meaningful rather than statistical noise, but larger cohorts will be needed to determine whether these response profiles are reproducible or prognostic.

### D. Relationship to Clinical Function and Disease Severity

The fatigue-change composite was not proportional to self-reported limb function. Clinical weakness and fatigue-related HD-sEMG change likely reflect overlapping but different aspects of disease. A muscle can be clinically weak because many motor units have been lost, but a less impaired muscle can still show unstable fatigue behavior if the remaining motor unit pool is heavily remodeled. Earlier or moderately affected muscles may retain enough motor unit reserve to generate variable compensatory responses, whereas more impaired muscles may have fewer remaining units and a narrower range of measurable fatigue-related change ^3,7,25–27,29,30^.

The severity-bin analysis was consistent with this interpretation. Composite values overlapped across severity groups, but the moderate impairment group showed the greatest variability and a modest shift toward more negative values. We view this as preliminary evidence that fatigue responses may be most variable when denervation, compensation, and emerging motor unit loss coexist. The sample is too small to claim a nonlinear disease trajectory, but the pattern supports physiological heterogeneity rather than a simple severity gradient.

### E. Future Directions

Larger ALS cohorts are needed to test whether the trend toward lower dispersion and the fatigue-response profiles observed here are reproducible. Future studies should include earlier disease stages, a broader range of muscles, and disease-control groups to determine whether these patterns are ALS-specific or reflect motor unit pathology more generally. Longitudinal sampling will be especially important because the most useful role for HD-sEMG may be tracking individualized physiological change over time, not classifying all ALS muscles with a single threshold.

Pairing HD-sEMG with structural measures could also improve interpretation. Ultrasound, for example, could test whether spatial activation patterns and fatigue-related signal changes correspond to muscle architecture, atrophy, or regional remodeling ^32,33^. Wearable HD-sEMG may eventually allow repeated sampling outside the clinic, where disease-related changes could be measured more often and under more natural conditions ^34,35^. Those tools would be most valuable if they preserve the muscle-level detail captured here while making longitudinal monitoring more practical.

## Conclusion

Baseline HD-sEMG structure was strongly spatial in both healthy and ALS muscles, and spatial features alone did not separate ALS from controls. Group separation emerged only when morphology and spatial features were analyzed together. Sustained contraction then shifted ALS toward a more variable fatigue-response profile involving RMS and spectral features. These findings support HD-sEMG as a tool for mapping neuromuscular organization and identifying physiological heterogeneity in ALS, rather than as a single uniform fatigue marker.

## Data Availability

All data produced in the present study are available upon reasonable request to the authors.

## Author Contributions

**Ernesto Bedoy:** Study design; Methodology (development of the HD-sEMG analytical framework, including feature domain selection, signal preprocessing, motor unit decomposition strategy, and principal component–based dimensionality reduction); Investigation (study piloting, experimental execution, and protocol implementation); Data acquisition (HD-sEMG recordings); Software (development of MATLAB analysis pipeline); Data curation; Formal analysis (motor unit decomposition, feature extraction, and statistical analysis); Visualization; Writing - original draft.

**Maia Brown:** Investigation (protocol approval, study piloting, and dynamometer application).

**Michael Christofidis:** Investigation (study piloting, study-session support); Data curation (processing of MUAP data using established analysis pipeline; preliminary exploration of spatial HD-sEMG decomposition workflows).

**Breanna Sullivan:** Investigation (participant recruitment, informed consent, dynamometer application, and data acquisition support); Clinical coordination.

**Tawfiq Al-lahham:** Clinical expertise; Methodology (clinical interpretation of EMG); Investigation (clinical demonstration of needle EMG and interpretation of findings; dynamometer training); Resources (clinical space coordination).

**Douglas Weber:** Conceptualization; Supervision.

**Christi Kolarcik:** Conceptualization; Funding acquisition; Supervision; Investigation (protocol approval and implementation, participant recruitment, consent, and study piloting); Clinical coordination.

All authors reviewed, edited, and approved the final manuscript.

## Data Availability

The data that support the findings of this study are available from the authors upon reasonable request, subject to institutional and ethical restrictions. Code and protocols will be made publicly available upon acceptance.

## Ethical Publication Statement

We confirm that we have read the Journal’s position on issues involved in ethical publication and affirm that this report is consistent with those guidelines.

## Acknowledgments

This work was supported by The ALS Association (Grant No. 24-SGP-723). The authors thank all study participants for their time and participation in this study. We also thank Max Gold for preliminary work exploring spatial HD-sEMG decomposition workflows, and Nikhil Verma, Max Murphy, Luigi Borda, Drew Beauchamp, and Prakarsh Yadav for technical guidance on decomposition methods. The authors also acknowledge the clinical staff at the University of Pittsburgh Medical Center (UPMC) for their assistance in coordinating clinical space for data collection. Finally, the authors are grateful to Destin Choice Route (JID; *God Does Like Ugly*) for inspiring the completion of this work done in honor of Carol Shaughnessy.

## Abbreviations

ALS: amyotrophic lateral sclerosis
ALSFRS-R: ALS Functional Rating Scale–Revised
BB: biceps brachii
DSDC: Decompose–Synthesize–Decompose–Compare
EMG: electromyography
FR: firing rate
FRV: firing rate variability
HD-sEMG: high-density surface electromyography
HF: high-frequency power
LF: low-frequency power
MUAP: motor unit action potential
MVC: maximum voluntary contraction
P2P: peak-to-peak amplitude
PC1: first principal component
PCA: principal component analysis
RMS: root-mean-square
SD: standard deviation
SE: standard error
SNR: signal-to-noise ratio
TA: tibialis anterior
UPMC: University of Pittsburgh Medical Center.

## References

1. Vasta R, De Mattei F, Tafaro S, Canosa A, Manera U, Grassano M, et al. Changes to Average Survival of Patients With Amyotrophic Lateral Sclerosis (1995–2018). Neurology: 2025;104:e213467. 10.1212/WNL.0000000000213467.

2. Moloney EB, de Winter F, Verhaagen J. ALS as a distal axonopathy: molecular mechanisms affecting neuromuscular junction stability in the presymptomatic stages of the disease. Front Neurosci: 2014;8. 10.3389/fnins.2014.00252.

3. Bekdik P, Baslo MB. Investigation of Ongoing Denervation and Reinnervation in Amyotrophic Lateral Sclerosis by Using Concentric Needle Electrode with Single Fiber Electromyography Method. Noro Psikiyatri Arsivi: 2023;60:298–303. 10.29399/npa.28162.

4. Keon M, Musrie B, Dinger M, Brennan SE, Santos J, Saksena NK. Destination Amyotrophic Lateral Sclerosis. Front Neurol: 2021;12. 10.3389/fneur.2021.596006.

5. Proudfoot M, Jones A, Talbot K, Al-Chalabi A, Turner MR. The ALSFRS as an outcome measure in therapeutic trials and its relationship to symptom onset. Amyotroph Lateral Scler Front Degener: 2016;17:414–425. 10.3109/21678421.2016.1140786.

6. Genge A, Cedarbaum JM, Shefner J, Chio A, Al-Chalabi A, Van Damme P, et al. The ALSFRS-R Summit: a global call to action on the use of the ALSFRS-R in ALS clinical trials. Amyotroph Lateral Scler Front Degener: 2024;25:382–387. 10.1080/21678421.2024.2320880.

7. de Carvalho M, Dengler R, Eisen A, England JD, Kaji R, Kimura J, et al. Electrodiagnostic criteria for diagnosis of ALS. Clin Neurophysiol Off J Int Fed Clin Neurophysiol: 2008;119:497–503. 10.1016/j.clinph.2007.09.143.

8. Turner MR, Kiernan MC, Leigh PN, Talbot K. Biomarkers in amyotrophic lateral sclerosis. Lancet Neurol: 2009;8:94–109. 10.1016/S1474-4422(08)70293-X.

9. Kirk EA, Sauerbrei BA. Accessing populations of motor units. eLife: 2024;13:e94764. 10.7554/eLife.94764.

10. Turner MR, Hardiman O, Benatar M, Brooks BR, Chio A, de Carvalho M, et al. Controversies and priorities in amyotrophic lateral sclerosis. Lancet Neurol: 2013;12:310–322. 10.1016/S1474-4422(13)70036-X.

11. Drost G, Stegeman DF, van Engelen BGM, Zwarts MJ. Clinical applications of high-density surface EMG: A systematic review. J Electromyogr Kinesiol: 2006;16:586–602. 10.1016/j.jelekin.2006.09.005.

12. Jordanic M, Rojas-Martínez M, Mañanas MA, Alonso JF. Spatial distribution of HD-EMG improves identification of task and force in patients with incomplete spinal cord injury. J NeuroEngineering Rehabil: 2016;13:41. 10.1186/s12984-016-0151-8.

13. McPherson LM, Negro F, Thompson CK, Sanchez L, Heckman C, Dewald J, et al. Properties of the Motor Unit Action Potential Shape in Proximal and Distal Muscles of the Upper Limb in Healthy and Post-Stroke Individuals. Conf Proc Annu Int Conf IEEE Eng Med Biol Soc IEEE Eng Med Biol Soc Annu Conf: 2016;2016:335–339. 10.1109/EMBC.2016.7590708.

14. Lara JE, Cheng LK, Rohrle O, Paskaranandavadivel N. Muscle-Specific High-Density Electromyography Arrays for Hand Gesture Classification. IEEE Trans Biomed Eng: 2022;69:1758–1766. 10.1109/TBME.2021.3131297.

15. Merletti R, Rainoldi A, Farina D. Myoelectric Manifestations of Muscle Fatigue. In: Electromyography: Physiology, Engineering, and Noninvasive Applications, Chap. 9. 2005. p. 233–258 10.1002/0471678384.ch9.

16. Nishikawa Y, Holobar A, Watanabe K, Takahashi T, Ueno H, Maeda N, et al. Detecting motor unit abnormalities in amyotrophic lateral sclerosis using high-density surface EMG. Clin Neurophysiol Off J Int Fed Clin Neurophysiol: 2022;142:262–272. 10.1016/j.clinph.2022.06.016.

17. Fernandes APM, Holanda LJ de, Lucena LC de, Silva KER da, Lopes ACSM, Borges DT, et al. Electromyography as a tool to motion analysis for people with Amyotrophic Lateral Sclerosis: A protocol for a systematic review. PLOS ONE: 2024;19:e0302479. 10.1371/journal.pone.0302479.

18. Donaghy R, Pioro EP. Neurophysiologic Innovations in ALS: Enhancing Diagnosis, Monitoring, and Treatment Evaluation. Brain Sci: 2024;14:1251. 10.3390/brainsci14121251.

19. Bashford J, Mills K, Shaw C. The evolving role of surface electromyography in amyotrophic lateral sclerosis: A systematic review. Clin Neurophysiol: 2020;131:942–950. 10.1016/j.clinph.2019.12.007.

20. Farina D, Leclerc F, Arendt-Nielsen L, Buttelli O, Madeleine P. The change in spatial distribution of upper trapezius muscle activity is correlated to contraction duration. J Electromyogr Kinesiol: 2008;18:16–25. 10.1016/j.jelekin.2006.08.005.

21. Rojas-Martínez M, Mañanas MA, Alonso JF. High-density surface EMG maps from upper-arm and forearm muscles. J NeuroEngineering Rehabil: 2012;9:85. 10.1186/1743-0003-9-85.

22. De la Fuente C, Weinstein A, Neira A, Valencia O, Cruz-Montecinos C, Silvestre R, et al. Biased instantaneous regional muscle activation maps: Embedded fuzzy topology and image feature analysis. Front Bioeng Biotechnol: 2022;10. 10.3389/fbioe.2022.934041.

23. Favretto MA, Cossul S, Andreis FR, Nakamura LR, Ronsoni MF, Tesfaye S, et al. Alterations of tibialis anterior muscle activation pattern in subjects with type 2 diabetes and diabetic peripheral neuropathy. Biomed Phys Eng Express: 2022;8. 10.1088/2057-1976/ac455b.

24. Lundsberg J, Björkman A, Malesevic N, Antfolk C. Inferring position of motor units from high-density surface EMG. Sci Rep: 2024;14:3858. 10.1038/s41598-024-54405-1.

25. Martineau É, Di Polo A, Vande Velde C, Robitaille R. Dynamic neuromuscular remodeling precedes motor-unit loss in a mouse model of ALS. Cleveland DW, Westbrook GL, editors. eLife: 2018;7:e41973. 10.7554/eLife.41973.

26. Henderson RD, McCombe PA. Assessment of Motor Units in Neuromuscular Disease. Neurotherapeutics: 2017;14:69–77. 10.1007/s13311-016-0473-z.

27. Duleep A, Shefner J. Electrodiagnosis of Motor Neuron Disease. Phys Med Rehabil Clin N Am: 2013;24:139–151. 10.1016/j.pmr.2012.08.022.

28. Sun J, Liu G, Sun Y, Lin K, Zhou Z, Cai J. Application of Surface Electromyography in Exercise Fatigue: A Review. Front Syst Neurosci: 2022;16. 10.3389/fnsys.2022.893275.

29. Enoka RM, Duchateau J. Muscle fatigue: what, why and how it influences muscle function. J Physiol: 2008;586:11–23. 10.1113/jphysiol.2007.139477.

30. de Carvalho M, Eisen A, Krieger C, Swash M. Motoneuron firing in amyotrophic lateral sclerosis (ALS). Front Hum Neurosci: 2014;8. 10.3389/fnhum.2014.00719.

31. Jahanmiri-Nezhad F, Hu X, Suresh NL, Rymer WZ, Zhou P. EMG-force relation in the first dorsal interosseous muscle of patients with amyotrophic lateral sclerosis. NeuroRehabilitation: 2014;35:307–314. 10.3233/NRE-141125.

32. Barnes SL, Simon NG. Clinical and research applications of neuromuscular ultrasound in amyotrophic lateral sclerosis. Degener Neurol Neuromuscul Dis: 2019;9:89–102. 10.2147/DNND.S215318.

33. Grimm A, Prell T, Décard BF, Schumacher U, Witte OW, Axer H, et al. Muscle ultrasonography as an additional diagnostic tool for the diagnosis of amyotrophic lateral sclerosis. Clin Neurophysiol: 2015;126:820–827. 10.1016/j.clinph.2014.06.052.

34. Garg R, Driscoll N, Shankar S, Hullfish T, Anselmino E, Iberite F, et al. Wearable High-Density MXene-Bioelectronics for Neuromuscular Diagnostics, Rehabilitation, and Assistive Technologies. Small Methods: 2023;7:e2201318. 10.1002/smtd.202201318.

35. Tacca N, Dunlap C, Donegan SP, Hardin JO, Meyers E, Darrow MJ, et al. Wearable high-density EMG sleeve for complex hand gesture classification and continuous joint angle estimation. Sci Rep: 2024;14:18564. 10.1038/s41598-024-64458-x.

