## Supplementary Information for "High-Density Surface Electromyography Reveals Shared Baseline Spatial Organization and Heterogeneous Fatigue Responses in Amyotrophic Lateral Sclerosis"

Intended for online-only publication.

### Supplementary Methods

#### High-Density Surface Electromyography (HD-sEMG) Acquisition and Preprocessing

Skin was prepared before electrode placement, and HD-sEMG was collected using separate decomposition and spatial recording systems. The Trigno Galileo sensor included four silver detection contacts (99.99% Ag) arranged with a 5 mm inter-electrode distance. The reference sensor was placed on the shoulder or calf, depending on the muscle tested. Signals were differentially amplified and digitized at the sensor with 16-bit resolution and a sampling frequency of 2222 Hz. Motor unit spike trains were extracted in NeuroMap software (v1.2.2, Delsys Inc.), which implements the Precision Decomposition III algorithm.

Decomposition recordings were screened first by signal quality. Recordings with signal-to-noise ratio (SNR) <3 dB were excluded, with SNR defined as electromyography (EMG) power during contraction relative to baseline noise power. After motor unit removal using the Decompose–Synthesize–Decompose–Compare (DSDC) test, motor units were retained only if decomposition accuracy was >90%. Additional post hoc filters excluded units with peak firing rate >60 Hz, mean firing rate <2 Hz, inter-pulse interval coefficient of variation >1.0, or peak motor unit action potential (MUAP) amplitude <20 µV. These criteria were used to remove units with implausibly high discharge rates, sparse firing, highly irregular discharge timing, or MUAPs near the noise floor.

Spatial HD-sEMG recordings were obtained with a textile grid of 32 Ag/AgCl electrodes arranged in a 4 × 8 configuration with an 8.75 mm inter-electrode distance. Reference and ground electrodes were placed on the elbow or knee depending on the muscle tested. Signals were recorded with a TMSi SAGA 32+ amplifier at 4000 Hz with 24-bit resolution. Conductive gel (Signa Gel, Parker, NJ) was applied to each electrode before placement.

Spatial grid signals were high-pass filtered with a fourth-order Butterworth filter at 5 Hz. Channels were flagged for review if they showed zero root-mean-square (RMS) or excessive RMS amplitude (>5 mV), abnormal deviation from the global common average reference (RMS greater than mean + 3 SD), or SNR <10 dB. Flagged channels were visually inspected with contraction windows overlaid. Channels were excluded only when noise, instability, saturation, dropout, or signal absence was confirmed. A longitudinal single-differential spatial filter was then applied along the limb to reduce far-field potentials and common-mode noise.

Contraction onset was determined retrospectively from the EMG envelope averaged across channels. The offset of the steady contraction phase was identified first, and the recorded contraction duration was then subtracted to estimate onset. This approach kept the analysis windows within the steady portion of each contraction and avoided including ramp-up or relaxation periods.

**Feature Computation**

Features were calculated at the channel or motor unit level and then collapsed to one value per observation, defined as Subject × Visit × Muscle × Target Force Level.

#### Fatigue Dynamics

Fatigue-dynamics features included RMS amplitude, low-frequency power (LF: 20–60 Hz), high-frequency power (HF: 170–350 Hz), motor unit firing rate (FR), and firing rate variability (FRV). RMS measured EMG amplitude in the time domain. LF and HF power captured frequency-domain changes associated with sustained contraction. FR and FRV were derived from decomposed motor unit spike trains and used to describe motor neuron discharge behavior and discharge stability.

RMS and spectral features were computed from the filtered spatial HD-sEMG signals. Spectral features were estimated with a Fast Fourier Transform and channel-level power spectral density. FR was calculated from inter-spike intervals, and FRV was calculated as the coefficient of variation of firing rate.

#### Motor Unit Morphology

Motor unit morphology was characterized using MUAP peak-to-peak amplitude, MUAP duration, and waveform complexity. Peak-to-peak amplitude was used as an amplitude-based MUAP measure. Duration and waveform complexity were included because collateral reinnervation and temporal dispersion can alter MUAP shape.

MUAP templates were derived by spike-triggered averaging of decomposed motor unit spike trains. Peak-to-peak amplitude was calculated as the difference between the maximum and minimum MUAP voltage. MUAP duration was measured from the initial deflection from baseline to the final return to baseline. Waveform complexity was quantified as the number of zero crossings in the MUAP waveform.

#### Propagation Features

Propagation features were estimated from the spatiotemporal evolution of MUAP waveforms across adjacent spatial HD-sEMG electrodes. These features included conduction velocity, temporal lag, and waveform correlation. Together, they were intended to capture conduction-related timing and waveform similarity across the grid. Conduction velocity was estimated from the temporal delay between MUAP peaks across neighboring channels. Lag represented the temporal shift between corresponding MUAP features across electrodes. Waveform correlation quantified similarity between spatially adjacent MUAP signals. Propagation estimates with poor model fit ($R^2 \leq 0.2$) were excluded.

Propagation features were not included in the final principal component analysis (PCA) models. In this dataset, conduction velocity and related lag or correlation estimates showed high variance and were sensitive to signal quality and propagation assumptions. Excluding these features prevented them from dominating the composite signal for technical rather than physiological reasons.

#### Spatial Organization

Spatial activation patterns were quantified from RMS activation maps. Spatial features included entropy, activation area, spatial dispersion, heterogeneity, cluster index, RMS variance, and peak-to-median RMS ratio. These measures described how activation was distributed across the electrode grid, including whether activity was broad, focal, clustered, or uneven. Activation maps were constructed from RMS amplitude at each electrode within each analysis window. Spatial entropy measured the uniformity of activation across the grid.

Activation area represented the proportion of electrodes exceeding the activation threshold. Dispersion measured the spread of activation relative to the RMS-map centroid. Heterogeneity and RMS variance captured channel-to-channel variability. Cluster index and peak-to-median ratio quantified localized activation and intensity gradients within the map.

**Data Aggregation and Normalization**

Two feature matrices were generated. The Early-window matrix used feature values from the first 10 s of steady contraction. The fatigue-change matrix expressed each feature as percent change from the Early window to the Late window, with the Late window defined as the final 10 s of contraction:

$$\Delta F_j(\%) \ = \ \frac{F_{j,Late} - F_{j,Early}}{F_{j,Early}} x100\%$$

where $F_{j,Early}$ and $F_{j,Late}$ are the values of feature j in the Early and Late analysis windows. Features were standardized before PCA because PCA is sensitive to feature scale. For group-specific PCA, features were standardized within each group so that loadings reflected the internal feature structure of Healthy or Amyotrophic Lateral Sclerosis (ALS) observations. For common-axis PCA, features were standardized to the Healthy control distribution so that Healthy and ALS observations could be projected onto the same axis while preserving group-level differences.

**Principal Component Analysis**

PCA was used to identify dominant patterns of HD-sEMG variation across feature domains. The first principal component (PC1) was used as the composite HD-sEMG signal because it explained the largest share of variance in each analysis. PC1 loadings were examined to determine which features and domains contributed most strongly to each composite.

Two PCA strategies were used. First, group-specific PCAs were run separately for Healthy Early, ALS Early, Healthy fatigue-change, and ALS fatigue-change observations. These analyses were used to describe the dominant loading structure within each group and condition. Because each group-specific PCA defines its own axis, these PC1 scores were not used for direct group comparison.

Second, common-axis PCAs were run across the full cohort for Early-window features and fatigue-change features. These analyses projected Healthy and ALS observations onto the same component, allowing group comparison of PC1 scores. Common-axis loadings were then examined to identify the feature domains that defined the shared axis.

The fatigue-change composite used for clinical coupling, severity-bin, and longitudinal analyses was defined as PC1 from the full-cohort PCA of all available Late-minus-Early percent-change features after Healthy-referenced standardization.

Domain-restricted follow-up analyses used the same common-axis PCA and mixed-effects modeling approach but were limited to features from one physiological domain at a time. These analyses were used to test whether group-level effects in the multifeature composite could be explained by a single feature domain.

**Statistical Analyses**

For common-axis group comparisons, linear mixed-effects models tested the effect of group on the PC1 score, with Healthy as the reference group. Contraction force and muscle were included as fixed effects when model structure allowed. Subject identity was included as a random intercept to account for repeated observations. In domain-restricted follow-up analyses, individual features were converted to Healthy-referenced z-scores so group effects could be interpreted relative to Healthy variability.

Cross-sectional relationships between the fatigue-change composite and functional impairment were evaluated in ALS observations using mixed-effects models with limb function score as a continuous predictor. Contraction force was included as a fixed effect, and subject identity was included as a random intercept. Severity-bin analyses compared the fatigue-change composite across mild, moderate, and severe impairment categories. These binned analyses were exploratory and were used to describe distributional patterns rather than fit a nonlinear disease model.

Longitudinal changes in the fatigue-change composite were assessed in participants with ALS using a mixed-effects model with time since baseline, limb function score, and contraction force as fixed effects and subject identity as a random effect. Feature stability in Healthy participants was evaluated with an analogous model restricted to Healthy observations. Longitudinal analyses were treated as exploratory because follow-up data were limited.

### Supplementary Results

#### Data Inclusion and Cohort Details

Baseline recordings were excluded from three participants: one participant with ALS and two Healthy participants. The first repeat visit from one participant with ALS was also excluded. These exclusions were due to poor EMG signal quality. One participant with ALS (ALS03) was unable to maintain continuous biceps brachii contractions and required multiple shorter contractions with concatenation or partial data selection. Because this differed from the standard sustained-contraction protocol, those recordings were excluded from the main analysis.

Follow-up data were available from five participants at Visit 2: two participants with ALS and three Healthy participants. Two participants with ALS completed a third visit. Additional recordings were excluded at the muscle, contraction-level, or electrode level because of poor EMG quality or unreliable motor unit decomposition. These exclusions are listed in **Table S5**.

**Maximum Voluntary Contraction (MVC) Strength**

MVC strength was lower in participants with ALS than in controls. For the biceps brachii, MVC was 40.4 ± 23.0 lbs in Healthy participants and 9.2 ± 5.9 lbs in participants with ALS (Mann–Whitney $p < 0.001$; **Figure S1**). For the tibialis anterior, MVC was 28.1 ± 14.5 lbs in Healthy participants and 13.0 ± 4.8 lbs in participants with ALS (Mann–Whitney $p = 0.003$; **Figure S1**). Longitudinal MVC values are shown in **Figure S1** and **Table S6.**

**Contraction Duration**

Contraction duration was shorter in ALS for the biceps brachii at 30% MVC (Healthy: 227.5 ± 59.4 s; ALS: 120.9 ± 89.0 s; Mann–Whitney $p < 0.05$; **Figure S2**). Differences at 50% MVC and in the tibialis anterior followed the same general direction but did not reach statistical significance. Longitudinal contraction durations are shown in **Figure S2** and **Table S7**.

**Decomposition Yield Consistency**

Motor unit decomposition yield did not differ significantly between groups across muscles or contraction intensities (Mann–Whitney $p > 0.10$). Yields overlapped between groups, ranging from 2 to 72 units in Healthy participants and from 6 to 58 units in participants with ALS. Low-yield recordings occurred in both groups, so low decomposition yield was not specific to ALS. Across visits, decomposition yield varied in both ALS (-20.4% to +183.3%) and Healthy participants (-87.5% to +138.1%) without a consistent ALS-specific decline.

**Representative Spatial Activation Maps**

Representative RMS activation maps were used to illustrate the type of spatial information captured by the HD-sEMG grid (**Figure 1D**). In the healthy example, activation appeared relatively distributed across the electrode grid, whereas the ALS example showed a more focal region of higher activation intensity. These examples demonstrate how spatial metrics such as entropy, dispersion, active area, heterogeneity, cluster index, and peak-to-median ratio quantify the distribution of muscle activity across the recording grid. These representative maps were not used to infer group-level spatial differences, which were tested quantitatively in the main analyses.

**Longitudinal Analysis and Healthy Stability**

Two participants with ALS completed three visits each, with an average interval of four months between visits. Clinical status was largely stable during follow-up. Consistent with this limited clinical change, no significant longitudinal progression of the fatigue-change composite was detected in ALS (slope = 0.155; $p = 0.250$; **Figure S3**). Healthy participants with repeat visits also showed no significant change in the fatigue-change composite over time ($\beta = 0.13$; $p = 0.238$; **Figure S3**). These results are useful as a stability check, but the sample is too small to draw conclusions about longitudinal progression.

**Supplementary Tables**

| Analysis | PC1 explained variance (%) | Spatial (%) | Fatigue (%) | Morphology (%) | Propagation (%) |
|---|---|---|---|---|---|
| ALS Early | 38.97 | 89.95 | 0.00 | 10.05 | 0.00 |
| ALS Fatigue-change | 31.74 | 57.91 | 34.36 | 7.73 | 0.00 |
| Healthy Early | 45.20 | 91.10 | 0.00 | 8.90 | 0.00 |
| Healthy Fatigue-change | 29.82 | 80.52 | 16.46 | 3.02 | 0.00 |

**Table S1. Group-specific principal component analysis summary.** Explained variance and percent of first principal component (PC1) loading weight contributed by each feature domain for the Healthy Early, ALS Early, Healthy fatigue-change, and ALS fatigue-change group-specific principal component analyses.

| Comparison | Cosine similarity | Spearman rho |
|---|---|---|
| Healthy Early vs ALS Early | 0.949 | 0.933 |
| Healthy Early vs Healthy Fatigue-change | 0.860 | 0.591 |
| ALS Early vs ALS Fatigue-change | 0.731 | 0.188 |
| Healthy Fatigue-change vs ALS Fatigue-change | 0.934 | 0.782 |

**Table S2. Loading-pattern similarity.** Pairwise similarity between PCA loading

| |
|---|
| patterns, quantified using cosine similarity and Spearman rank correlation of absolute loadings. |

| Analysis | ALS minus Healthy estimate | SE | t-statistic | df | p-value |
|---|---|---|---|---|---|
| Baseline multifeature common-axis | -1.372 | 0.420 | -3.265 | 89 | 0.0016 |
| Fatigue-change common-axis | -0.614 | 0.570 | -1.079 | 89 | 0.2837 |
| Baseline spatial-only common-axis | 0.341 | 0.457 | 0.746 | 81 | 0.4579 |

**Table S3. Common-axis principal component analysis group comparisons.**

Mixed-effects model estimates for ALS versus Healthy group comparisons using the baseline multifeature common-axis PCA, baseline spatial-only common-axis PCA, and fatigue-change common-axis PCA.

| Feature | Healthy mean | ALS mean | ALS minus Healthy | raw β, z-score units | p-value | Direction |
|---|---|---|---|---|---|---|
| Dispersion | 2.268 | 2.206 | -0.062 | -0.765 | 0.0573 | ALS lower |
| Spatial Entropy | 3.168 | 3.128 | -0.041 | -0.540 | 0.2130 | ALS lower |
| Peak-to-Medi | 2.118 | 1.887 | -0.231 | -0.177 | 0.2594 | ALS lower |

| | | | | | | |
|---|---|---|---|---|---|---|
| an Ratio | | | | | | |
| Active Area | 0.652 | 0.603 | -0.049 | -0.193 | 0.4185 | ALS lower |
| Spatial Kurtosis | 2.965 | 2.920 | -0.045 | -0.154 | 0.6375 | ALS lower |
| Cluster Index | 1.332 | 1.307 | -0.025 | -0.067 | 0.6732 | ALS lower |
| RMS Variance | 1071.305 | 1098.581 | 27.277 | -0.059 | 0.8306 | ALS lower |
| Heterogeneity | 0.396 | 0.402 | 0.006 | 0.012 | 0.9480 | ALS higher |

**Table S4. Individual Early-window spatial feature models.** Mixed-effects model results for each Early-window spatial feature. Group effects are shown as ALS minus Healthy. Standardized group effects are reported in Healthy-referenced z-score units.

| **Visit 1 (Baseline)** | | | |
|---|---|---|---|
| **Subject ID** | **Muscle** | **% MVC** | **Electrode** |
| H01 | BB | 30 | Decomposition |
| H01 | TA | 30/50 | Spatial |
| H02 | TA | 50 | Decomposition |
| H03 | BB | 30/50 | Spatial |
| H04 | BB | 30 | Decomposition |
| H05 | BB | 50 | Decomposition |
| H05 | TA | 50 | Decomposition |
| H07 | TA | 30/50 | Decomposition |
| H10 | BB | 50 | Decomposition |
| ALS01 | TA | 30 | Decomposition |

| | | | |
|---|---|---|---|
| ALS02 | BB & TA | 30/50 | Spatial |
| ALS02 | BB | 30 | Decomposition |
| ALS04 | BB | 30 | Decomposition |
| ALS05 | BB | 30/50 | Decomposition |
| **Visit 2** | | | |
| **Subject ID** | **Muscle** | **% MVC** | **Electrode** |
| H03 | BB | 50 | Spatial |
| ALS02 | TA | 30/50 | Decomposition |
| **Visit 3** | | | |
| **Subject ID** | **Muscle** | **% MVC** | **Electrode** |
| ALS01 | BB | 30 | Decomposition |

**Table S5. Data exclusions.** Data from 17 participants were included in the analysis. The table lists muscles, contraction levels expressed as percent maximum voluntary contraction (MVC), and electrode recordings excluded because of poor EMG signal quality or unreliable motor unit decomposition. Decomposition refers to the 4-channel decomposition recordings, and Spatial refers to the 32-channel HD-sEMG grid recordings. Overall, 16.7% of Decomposition data and 7.3% of Spatial data were excluded after quality control.

| **Biceps Brachii** | | | | |
|---|---|---|---|---|
| **Subject** | **Visit 1** | **Visit 2** | **Visit 3** | **% Change** |
| H02 | 27 lbs | 33 lbs | NA | 22.2 % |
| H03 | 33.2 lbs | 42 lbs | NA | 26.5% |
| H05 | 82.4 lbs | 57.1 lbs | NA | -30.7% |

| | | | | |
|---|---|---|---|---|
| ALS01 | 18 lbs | 19 lbs | 25 lbs | 38.9% |
| ALS02 | 15.4 lbs | 22.2 lbs | 22.1 lbs | 43.5% |
| ALS05 | 5.5 lbs | 3.7 lbs | NA | -32.7% |
| **Tibialis Anterior** | | | | |
| H02 | 22.8 lbs | 31.6 lbs | NA | 38.6% |
| H03 | 29.3 lbs | 28.9 lbs | NA | -1.4% |
| H05 | 50.0 lbs | 34.6 lbs | NA | -30.8% |
| ALS01 | 15.9 lbs | 12.2 lbs | 12.5 lbs | -21.4% |
| ALS02 | 20.9 lbs | 28.4 lbs | 38.1 lbs | 82.3% |
| ALS05 | 11.5 lbs | 6.4 lbs | NA | -44.3% |

**Table S6. Longitudinal changes in maximum voluntary contraction (MVC).** MVC values (lbs) and percent change between the first and last available visit for each subject. Biceps brachii (BB) results are shown in the top panel and tibialis anterior (TA) results in the bottom panel.

| **Biceps Brachii** | | | | |
|---|---|---|---|---|
| **Subject** | **Visit 1 (30/50% MVC)** | **Visit 2 (30/50% MVC)** | **Visit 3 (30/50% MVC)** | **% Change (30/50% MVC)** |
| H02 | 300/150 s | 230/135 s | NA | -23.3%/-10.0% |
| H03 | 260/60 s | 60/73 s | NA | -76.9%/21.7% |
| H05 | 180/40 s | 165/85 s | NA | -8.3%/112.5% |
| ALS01 | 240/180 s | 300/300 s | 300/240 s | 25%/33.3% |
| ALS02 | 240/180 s | 200/70 s | 220/97 s | -8.3%/-46.1% |
| ALS05 | 120/75 s | 53/57 s | NA | -55.8%/-24.0% |
| **Tibialis Anterior** | | | | |

| | | | | |
|---|---|---|---|---|
| H02 | 300/200 s | 270/150 s | NA | -10%/-25% |
| H03 | 210/180 s | 300/115 s | NA | 42.9%/-36.1% |
| H05 | 210/100 s | 215/150 s | NA | 2.4%/50% |
| ALS01 | 240/180 s | 300/300 s | 300/300 s | 25%/66.7% |
| ALS02 | 300/300 s | 300/300 s | 300/260 s | 0%/-13.3% |
| ALS05 | 270/100 s | 60/67 s | NA | -77.8%/-33.0% |

**Table S7. Longitudinal changes in contraction duration.** Contraction durations (seconds) and percent change between the first and final visit for each subject. Biceps brachii (BB) results are shown in the top panel and tibialis anterior (TA) results in the bottom panel.

#### Supplementary Figure

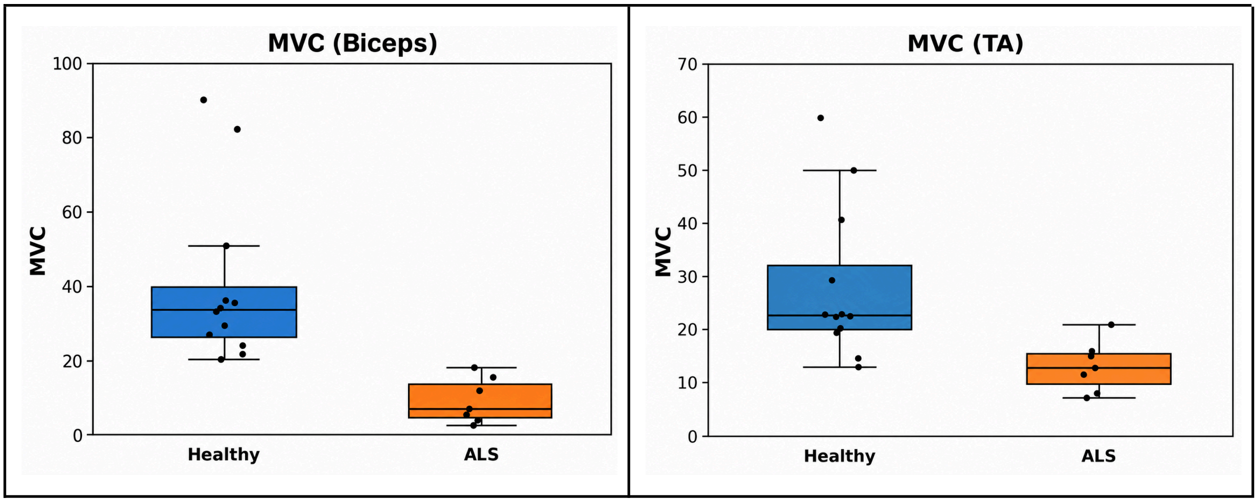

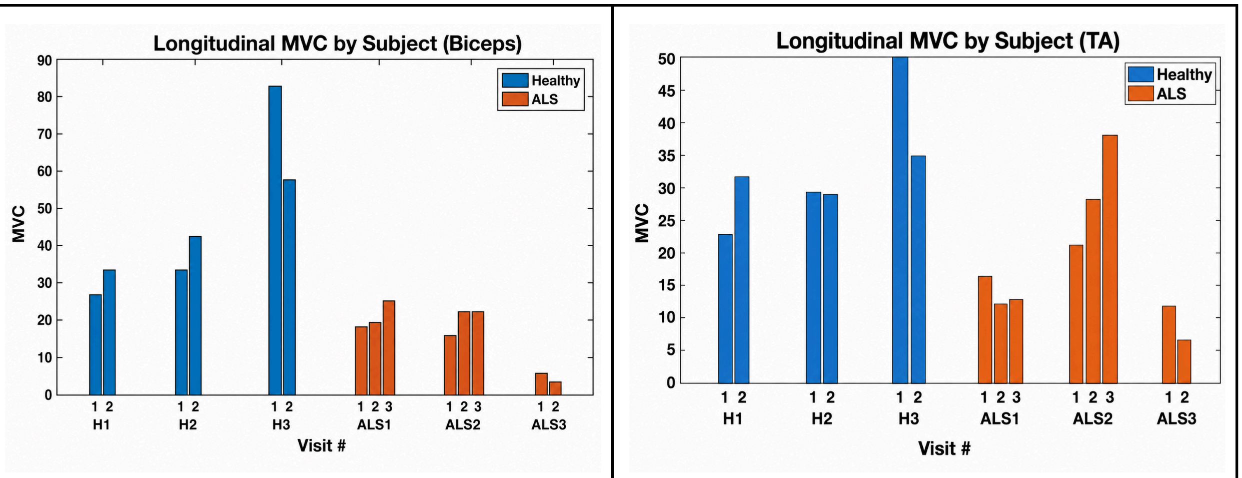


**Figure S1. Maximum voluntary contraction (MVC) strength.** Absolute MVC values (lbs) are shown for the biceps brachii (Biceps, left column) and tibialis anterior (TA, right column). Baseline group comparisons are shown in the top row, with individual participant values overlaid on boxplots for Healthy and ALS groups. Longitudinal MVC values are shown in the bottom row for participants with repeat visits, with bars representing MVC strength at each visit for individual subjects. Healthy participants are shown in blue and participants with ALS are shown in orange.

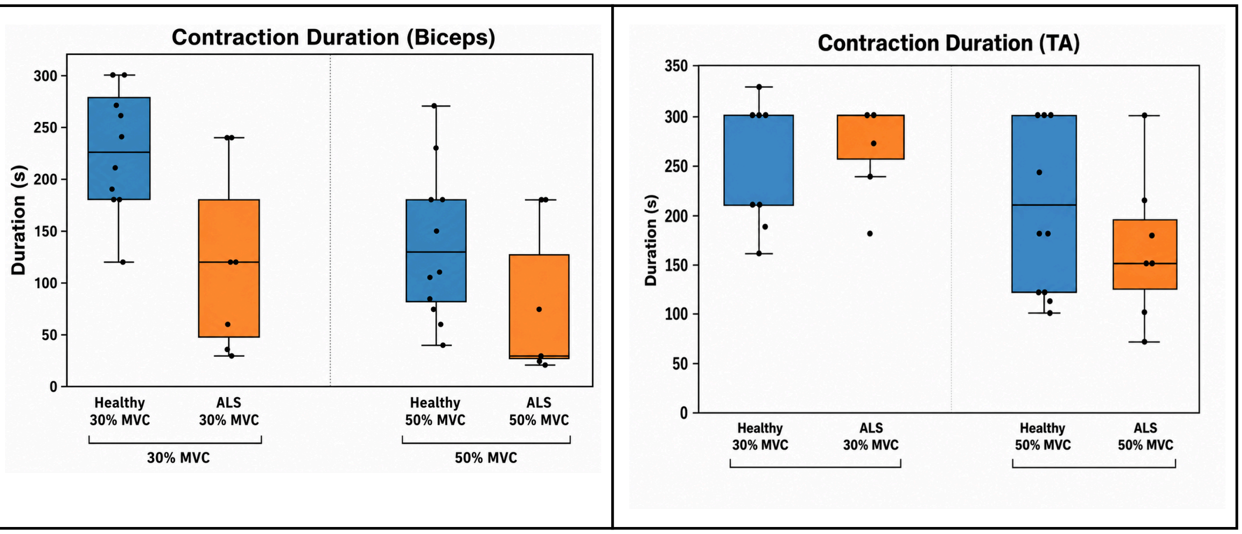

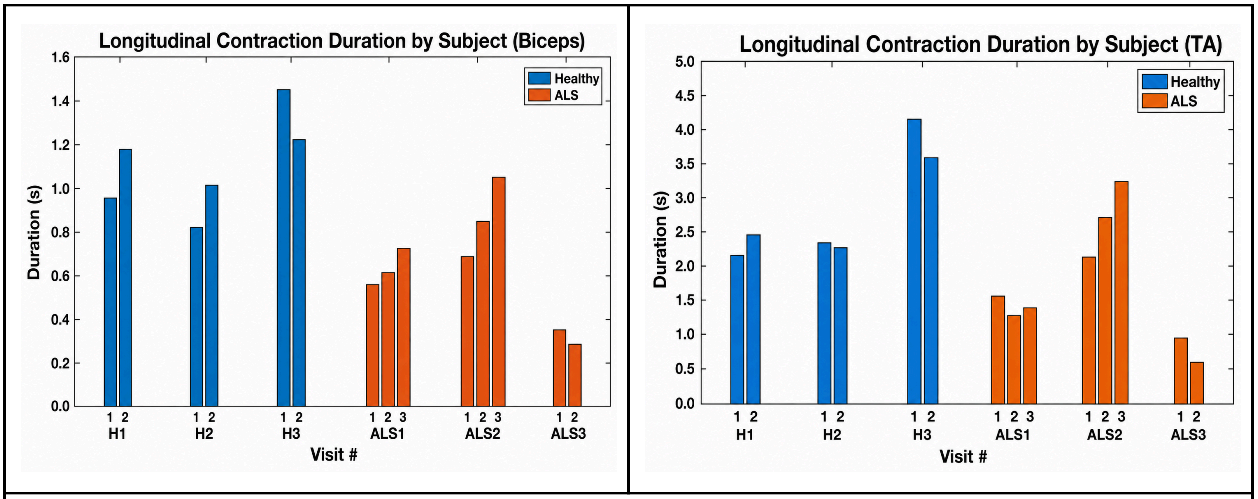


**Figure S2. Duration of sustained contractions.** Steady contraction durations (s) are shown for the biceps brachii (Biceps, left column) and tibialis anterior (TA, right column). Baseline group comparisons are shown in the top row, with individual participant values overlaid on boxplots for Healthy and ALS groups at 30% and 50% MVC. Longitudinal contraction durations are shown in the bottom row for participants with repeat visits, with bars representing contraction duration at each visit for individual subjects. Healthy participants are shown in blue and participants with ALS are shown in orange.

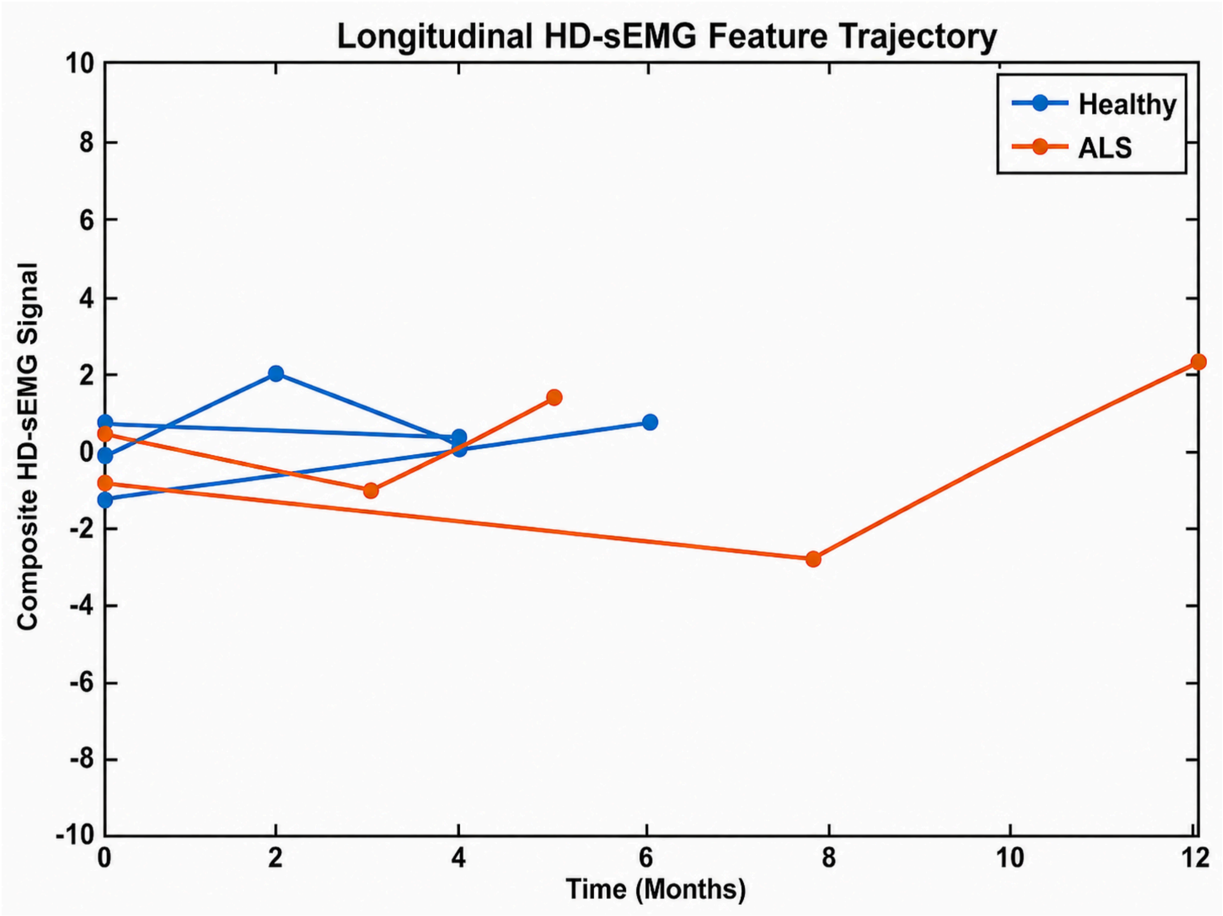


**Figure S3. Longitudinal composite HD-sEMG feature trajectories.** Composite HD-sEMG feature values are shown over time for participants with repeated visits. Each line represents one participant, with values averaged across muscles and contraction intensities at each visit. Healthy participants are shown in blue, and participants with ALS are shown in red. Healthy participants showed stable feature values over time. Participants with ALS showed minimal longitudinal change, consistent with largely stable clinical status during follow-up.
